# Detection of *Hepatovirus A* (HAV) in wastewater indicates widespread national distribution and association with socioeconomic indicators of vulnerability

**DOI:** 10.1101/2024.07.28.24311142

**Authors:** Alessandro Zulli, Elana M. G. Chan, Alexandria B. Boehm

## Abstract

Wastewater-based epidemiology, which seeks to assess disease occurrence in communities through measurements of infectious disease biomarkers in wastewater, may represent a valuable tool for understanding occurrence of hepatitis A infections in communities. In this study, we measured concentrations of *Hepatovirus A* (HAV) RNA, in samples from 191 wastewater treatment plants spanning 40 US states and the District of Columbia from September 2023 to June 2024 and compared the measurements with traditional measures of disease occurrence. Nationally, 13.76% of the 21,602 wastewater samples were positive for HAV RNA, and both concentrations and positivity rates were associated with NNDSS hepatitis A case data nationally (Kendall rank correlation coefficient = 0.20, concentrations; and 0.33, positivity rate; both p<0.05). We further demonstrated that higher rates of wastewater HAV detection were positively associated with socioeconomic indicators of vulnerability including homelessness and drug overdose deaths (both p<0.0001). Areas with above average levels of homelessness were 48% more likely to have HAV wastewater detections, while areas with above average levels of drug overdose deaths were 14% more likely to have HAV wastewater detections. Using more granular case data, we present a case study in the state of Maine that reinforces these results and suggests a potential lead time for wastewater over clinical case detection and exposure events. The ability to detect HAV RNA in wastewater before clinical cases emerge could allow public health officials to implement targeted interventions like vaccination campaigns.

**Importance:** Despite the existence of a highly effective vaccine for Hepatitis A, outbreaks in vulnerable populations remain common. The disease can be asymptomatic or subclinical, and disproportionately impacts populations with inadequate access to healthcare, leading to a severe underestimation of the occurrence of this viral infection. This study investigates the potential for wastewater measurements of biomarkers of the causative agent of hepatitis A (HAV RNA) to provide insights into disease occurrence. Results highlight the potential for wastewater-based epidemiology to be a complementary tool to traditional surveillance for monitoring and controlling HAV transmission.

## Introduction

Hepatitis A outbreaks have rapidly become a persistent and severe public health threat in the United States amongst vulnerable populations such as men who have sex with men, persons who use drugs and persons experiencing homelessness.^1–3^ Hepatitis A is a contagious liver infection caused by the *Hepatovirus A* virus (HAV) and transmitted through person-to-person contact or by ingesting contaminated food and water.^4,5^ Globally, the virus is particularly common in low and middle income countries, with 90% of children in these areas having been infected by age 10.^6^ Annually, HAV is responsible for an estimated 39,000 deaths, 1.4 million symptomatic infections, and 158.9 million total infections worldwide.^7,8^ Despite the virus’s worldwide ubiquity, populations in high income countries with high levels of access to clean water and improve sanitation, such as the United States (US), are susceptible to HAV due to a lack of childhood exposure and/or vaccination (74% susceptible).^5,9^ This is of particular concern in recent years as despite the availability of an effective vaccine and well-understood hygiene-based prevention measures, infections have increased in high-income countries, exacerbated by socioeconomic factors and increasing globalization.^1,5,9–11^

The introduction of the HAV vaccine in 1996 led to a 95% decrease in annual cases in the US by 2011.^11^ Despite these successful mitigation efforts, cases began to rise again in 2012, and outbreaks began to be recognized nationwide in 2016.^11^ Between 2016 and the present (2024), 37 states have suffered outbreaks of the virus, resulting in 44,926 cases, 27,457 hospitalizations, and 424 deaths.^3^ These outbreaks have largely occurred within vulnerable populations of persons reporting drug use and experiencing homelessness, who are often unvaccinated.^11,12^ While hepatitis A has been a nationally notifiable disease since 1966, it is often asymptomatic and affects populations with inadequate access to healthcare, leading to a severe underestimation of the occurrence of this viral infection.^1,2,10,13^ This has lead to increasing interest in alternative methods of monitoring such as wastewater.^14^

HAV infection presents clinically with several gastrointestinal symptoms including nausea, vomiting, diarrhea, and dark-coloured urine.^5,6,11^ HAV is mainly transmitted through person-to-person contact or the ingestion of contaminated food or water.^4,5^ Previous studies have shown HAV detection in stool and urine, with concentrations in stool being similar to serum levels of the virus.^15–18^ Further, HAV has been shown to be highly prevalent in wastewater, with a recent meta-analysis reporting 31.4% of all wastewater samples positive for the virus.^14,19,20^ A study in Cordoba, Argentina, demonstrated that the percentage of wastewater samples positive for HAV coincided with known outbreaks.^14^ These findings, along with known gaps in detection of HAV in vulnerable populations, makes HAV an ideal candidate for the implementation of wastewater monitoring.^9,11,12^

In this study, we measured concentrations of HAV genomic RNA in samples from 191 wastewater treatment plants spanning 40 U.S. states and the District of Columbia from 11 September 2023 to 1 June 2024 and compared the measurements with traditional measures of disease occurrence. We demonstrate that detections and concentrations of HAV RNA in wastewater are associated with incident HAV cases in the population, compare the frequency of detection of HAV in wastewater to the prevalence of known risk-factors such as drug use and homelessness, and present a detailed case study of a known outbreak in Maine. Together, the findings demonstrate the potential for using wastewater surveillance to inform public health interventions.

## Methods

This study was reviewed by the Stanford University Ethical Review Board (IRB) and determined that this project does not meet the definition of human subject research as defined in federal regulations 45 CFR 46.102 or 21 CFR 50.3.

### Wastewater data: sample collection

Wastewater measurements were made prospectively as part of an ongoing wastewater monitoring program. Between 11 September 2023 and 1 June 2024, wastewater samples (either 24-hour composited influent or grab samples from the primary clarifier) were collected by wastewater treatment plant staff. Samples were typically obtained three times per week, but as frequently as daily, and shipped overnight to the laboratory at 4°C and processed immediately upon receipt with no storage. Samples were collected from 191 distinct wastewater treatment plants (WWTPs) across a total of 40 US states and the District of Columbia (Table S1) for a total of 21,471 samples.

### Wastewater data: pre-analytical processing

Several studies have demonstrated that viruses, including HAV, partition favorably to the solid fraction of wastewater.^18,21,22^ As a result, HAV nucleic acids were measured in the solid phase of wastewater for this project. Details of the specific wastewater solid isolation protocols and nucleic acid extraction methods are presented in Boehm et al.^23^ In short, solids were isolated (dewatered) from samples by centrifugation and then an aliquot was suspended at 75 mg/ml in bovine coronavirus vaccine (BCoV) spiked buffer. After homogenization and additional centrifugation, the 300 µl of supernatant was input into a commercial nucleic acid extraction kit, and then an inhibitor removal kit; the resultant purified nucleic-acid extract was 50 µl. Negative extraction controls consisted of BCoV spiked into buffer. Nucleic acids were extracted from 6 or 10 (Table S1) replicate aliquots of each sample and used immediately as template (no storage) in ddRT-PCR, as described below. The dry weight of the solids were determined using oven drying using an additional aliquot of dewatered solids.^23^

### Wastewater data: analytical processing

Droplet digital reverse transcription polymerase chain reaction (ddRT-PCR) was used to measure concentrations of nucleic acid targets. The HAV assay is the assay originally published by Jothikumar et al. that targets the 5’ untranslated region (UTR) of the HAV RNA genome.^24^ Those authors thoroughly confirmed the assay sensitivity and specificity for HAV virus. The assay was run in multiplex using a probe-mixing approach. The other assays that were multiplexed included those targeting influenza A and B virus, the N gene of SARS-CoV-2, respiratory syncytial virus, norovirus GII, and rotavirus; results for these assays are not provided herein. Experiments showed that the multiplexed assays do not interfere with each other (see SI and Figure S1). Pepper mild mottle virus (PMMoV) was also measured as an endogenous positive control and BCoV was used as a spiked in control; these assays were run in duplex as described elsewhere.^25^ Each of the replicate nucleic-acid extracts was run in its own well to measure HAV (6 or 10 wells per sample) and 2 randomly chosen extracts were run in 2 wells to measure PMMoV and BCoV. The exception was the samples for which 10 replicate extracts were available; for these, each of the 10 replicates were run in their own well to measure PMMoV and BCoV (Table S1). Samples were run on 96 well plates, and each plate contained one well consisting of a positive PCR control, two no template controls, and two negative extraction controls. The positive control for HAV was synthetic cDNA (ATCC VR-3257SD), positive controls for PMMoV and BCoV are described elsewhere.^25^

ddRT-PCR was performed on 20 µl samples from a 22 µl reaction volume, prepared using 5.5 µl template, mixed with 5.5 µl of One-Step RT-ddPCR Advanced Kit for Probes (Bio-Rad 1863021), 2.2 µl of 200 U/µl Reverse Transcriptase, 1.1 µl of 300 mM dithiothreitol (DTT) and primers and probes mixtures at a final concentration of 900 nM and 250 nM respectively. Primer and probes for assays were purchased from Integrated DNA Technologies (IDT, San Diego, CA) (Table 2). HAV was measured in reactions using undiluted template whereas PMMoV and BCoV were measured using template diluted 1:100 in molecular grade water.

Droplets were generated using the AutoDG Automated Droplet Generator (Bio-Rad, Hercules, CA). PCR was performed using Mastercycler Pro (Eppendforf, Enfield, CT) with the following cycling conditions: reverse transcription at 50°C for 60 minutes, enzyme activation at 95°C for 5 minutes, 40 cycles of denaturation at 95°C for 30 seconds and annealing and extension at 59°C (for HAV) or 56°C (for PMMoV and BCoV) for 30 seconds, enzyme deactivation at 98°C for 10 minutes then an indefinite hold at 4°C. The ramp rate for temperature changes were set to 2°C/second and the final hold at 4°C was performed for a minimum of 30 minutes to allow the droplets to stabilize. Droplets were analyzed using the QX200 (PMMoV/BCoV) or the QX600 Droplet Reader (HAV) (Bio-Rad). A well had to have over 10,000 droplets for inclusion in the analysis. All liquid transfers were performed using the Agilent Bravo (Agilent Technologies, Santa Clara, CA).

Thresholding was done using QuantaSoft™ Analysis Pro Software (Bio-Rad, version 1.0.596) and QX Manager Software (Bio-Rad, version 2.0). Replicate wells were merged for analysis of each sample. In order for a sample to be recorded as positive, it had to have at least 3 positive droplets.

Concentrations of RNA targets were converted to concentrations in units of copies (cp)/g dry weight using dimensional analysis.^26^ The error is reported as standard deviations and includes the errors associated with the Poisson distribution and the variability among the replicate wells. Three positive droplets across merged wells corresponds to a concentration between ∼500-1000 cp/g; the range in values is a result of the range in the equivalent mass of dry solids added to the wells and the number of wells (6 or 10). Data collected as part of the study are available from the Stanford Digital Repository (https://purl.stanford.edu/qf850cv6453).

### Clinical surveillance data

The National Notifiable Disease Surveillance System (NNDSS) is a nationwide collaboration to which health departments share health information about nationally notifiable infectious and noninfectious diseases.^27^ Total case reports are compiled on a weekly basis using data from 50 states, the District of Columbia, New York City, and 5 territories. For this study, we used publicly available information on NNDSS for laboratory-confirmed HAV incident cases from 1 July, 2023 to 1 June, 2024. Dates of case determination varied but included the following in order of preference: date of disease start, date of diagnosis, date of laboratory result, date of first report to public health system, or date of state report.^28^ This case data were adjusted for population based on data from the US census and converted to a rate of cases per 100,000 population.^29^ While this dataset is limited to facilities which report cases to the NNDSS system, it is the most complete dataset available for comparisons across the entire United States.

For a more focused comparison with potentially more timely and time-resolved data, we conducted an in depth analysis using data from the state of Maine. The state of Maine was identified as of particular interest due to a high occurrence of the HAV nucleic acid targets in wastewater (described in results), a high rate of clinical cases and known outbreaks during the study period.^1,3,30^ Case data for the state of Maine were provided through the Division of Disease Surveillance in the Maine Department of Health Human Services (MDPH), which is available upon request.^31^ The data are provided as the first positive test for each de-identified individual by date of specimen collection. This de-identified data were reported by date of specimen collection.

### Homelessness and drug overdose data

Data about individuals experiencing homelessness was aggregated from the US Department of Housing and Urban Development and the 2023 Annual Homeless Assessment Report to Congress.^32,33^ Rates are reported as unhoused persons per 10,000 population at the state level. Drug overdose data were aggregated from the CDC WONDER database and reported as the number of deaths per 100,000 population at the state level.^34,35^ Data on individuals experiencing homelessness and drug overdoses was available as yearly averages on a state level basis for 2023 at the latest and is assumed to be representative of values during our study period (2023-2024).

### Data analysis

We ran several comparisons between summary statistics of HAV concentrations in wastewater and clinical surveillance case rates (cases per 100,000/week). Case data from NNDSS, available on a weekly basis, was used by converting cases to cases per 100,000 people for each state. First, we ran comparisons between weekly wastewater concentrations and weekly clinical cases aggregated at the national and state level. Non-detect results in wastewater were treated as 0 copies per gram dry weight (cp/g). Weekly wastewater concentrations were calculated by taking the Morbidity and Mortality Weekly Report (MMWR) weekly average concentrations at each WWTP and adjusting their contributions to the state total as previously described by population (SI Eq 1).^36^ We then calculated the weekly percentage of positive detections of wastewater HAV by counting all positive observations for a single WWTP and dividing by the total number of observations in each week. This process was repeated by aggregating data on a state basis. All weekly data were aggregated by MMWR week. Kendall’s tau rank correlation coefficients were used to assess the association between weekly NNDSS case rates and both average wastewater concentrations and percentage positive detections at both the state and national levels.

We also assessed the relationship between the percentage of positive wastewater detections of HAV and indicators of homelessness and drug overdose deaths.^14^ Percentage of positive wastewater detections was used in accordance with previous studies.^14^ For this analysis, the percent of positive wastewater detections over the entire study period was calculated for each state. We then separated the states into two groups; one group with above or equal to the mean and one group with below mean metrics of individuals experiencing homelessness, and also drug overdose deaths. We then calculated the mean percentage of positive wastewater detections in each group across the study period. Statistical significance and 95% confidence intervals were calculated through bootstrapping.^37^ The null hypothesis tested was that there was no significant difference between the percentage of wastewater detections in the two groups of observations. For each variable and group, a distribution of values was generated through random sampling with replacement 30,000 times.

We further present an in-depth case study comparing wastewater HAV nucleic acid concentrations and new HAV diagnoses in the state of Maine. In addition to the analyses described above, we calculated Kendall’s tau rank correlation coefficient between monthly average concentrations and monthly diagnoses, and present cross-correlation data to examine whether wastewater HAV data lead or lag diagnoses. We focused on two specific counties with identified hepatitis A outbreaks for which the communities were served by WWTPs participating in the study: Androscoggin and Cumberland. Case data were used in conjunction to identify these counties as having a high likelihood of HAV transmission. Kendall’s rank correlation coefficient was used for cross-correlation analysis on a weekly basis.

All statistical analyses were performed in RStudio using R Statistical Software (version 4.3.2).^38^ The p-value threshold to assign significance was 0.05. Monthly average concentrations were calculated using simple averages, and cross-correlation was assessed using Kendall’s rank correlation coefficient and the ccf() function from the t-series package (version 0.10-56).

## Results

### QA/QC

Results are reported following the Environmental Microbiology Minimal Information (EMMI) guidelines (Figure S2). All positive and negative controls were positive and negative, respectively. Median (IQR) BCoV recoveries across all wastewater samples were 1.07 (0.81, 1.42) indicating good recovery across all samples. Recoveries exceeding 1 are the result of uncertainties in the measurement of BCoV added to the buffer matrix. PMMoV levels were elevated in all samples indicating lack of gross extraction failures (median = 4.6×10^8^ cp/g, min = 5.0×10^5^ cp/g, max = 2.02×10^11^ cp/g, IQR= 1.8×10^8^ - 6.2×10^8^).

### National overview

The study period ran from 11 September 2023 to 1 June 2024 and spanned 191 distinct WWTPs in 40 states and the District of Columbia (Table S1). States had between 1 and 57 treatment plants enrolled. Population coverage of the sewersheds as a function of total state population ranged from 0.13% to 59.5% of the population of each state (median: 5.75%). Each WWTP provided between 31 to 275 samples during the study period (median=113). In total, 21,602 samples were collected and analyzed across all WWTPs. HAV RNA concentrations ranged from below the limit of detection (approximately 1000 cp/g) to ∼12,000,000 cp/g, with 13.8% of all samples having detectable HAV RNA. Of the 40 states included in the study, 38 of them and the District of Columbia had at least one positive HAV detection (Mississippi and Washington state had no positive detections), and of the 191 WWTPs, 147 had at least one positive HAV detection (77%). The percentage of positive samples in WWTPs with at least one detection ranged between 0.9% to 95.1%. Table S2 provides a summary of percentage positive detections at each WWTP. Detections of HAV RNA varied widely by location. The highest percentage of detections was found in Portland Water District in Maine (93.47%), while the lowest levels of detection were 0% in 43 WWTPs. At the state level, the highest levels of detection were found in Massachusetts, Kansas, and Maine (45.1%, 37.9%, 34.27%) while the lowest levels of detection were in Washington and Mississippi (0%). Nationally, 13.76% of all samples were positive for HAV RNA.

Case rates from the NNDSS were compared to population weighted wastewater concentrations on a statewide basis. Table 1 shows Kendall’s tau for each state. Five (of 40) states had positive and significant correlations (p<0.05). States marked with NA (7 states total) had either no detectable HAV in wastewater or 0 recorded HAV cases, leading to no rank-correlation coefficient being calculated. For the remaining 28 states, the rank correlation was not statistically different from 0.

**Table 1.**
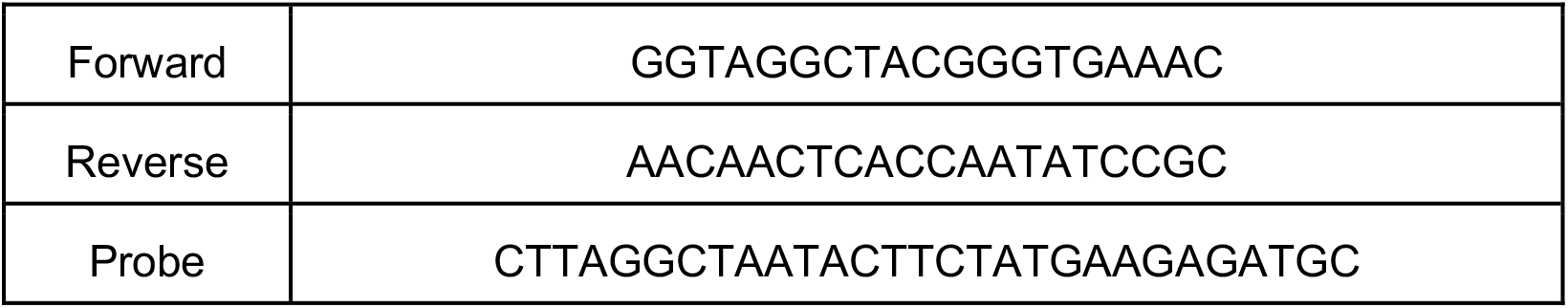
Primers and probes used for detection of *Hepatovirus A* (HAV) nucleic acids, published and validated by Jothikumar et al.^24^ Primers and probes were purchased from Integrated DNA Technologies (Coralville, IA, USA). The probes contained fluorescent molecule FAM and quenchers (5′ FAM/ZEN/3′ IBFQ); FAM, fluorescein amidite; ZEN, a proprietary internal quencher from Integrated DNA Technologies (Coralville, IA, USA); and IBFQ, Iowa Black FQ. Amplicon size is 89 base pairs.

**Table 2:**
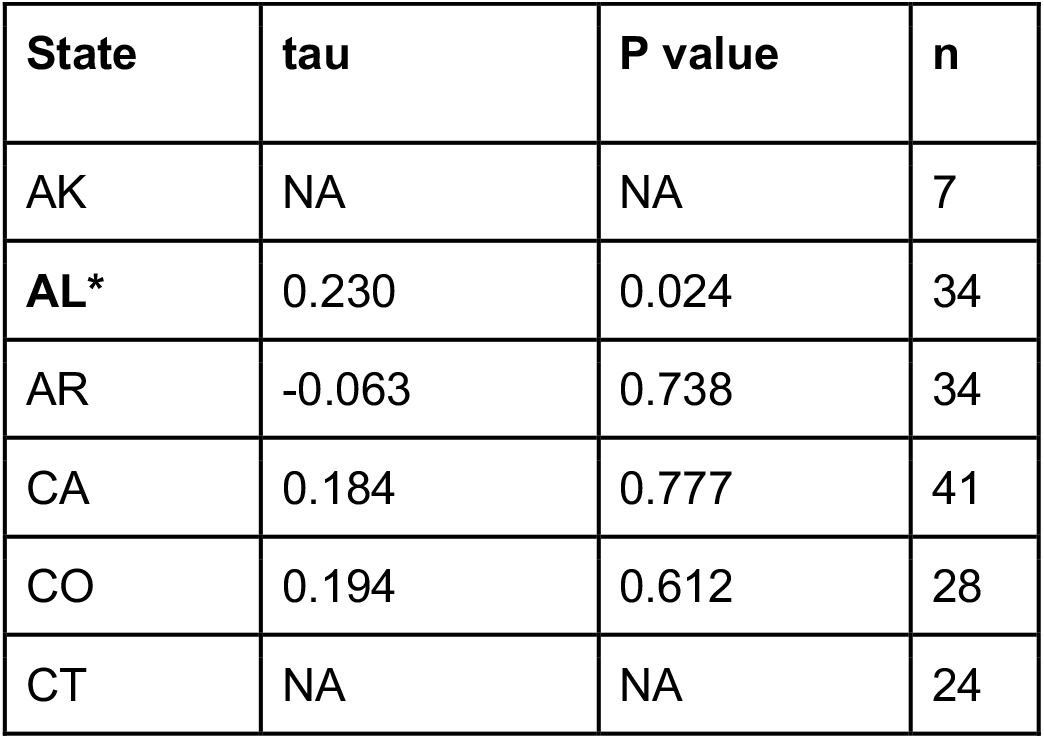
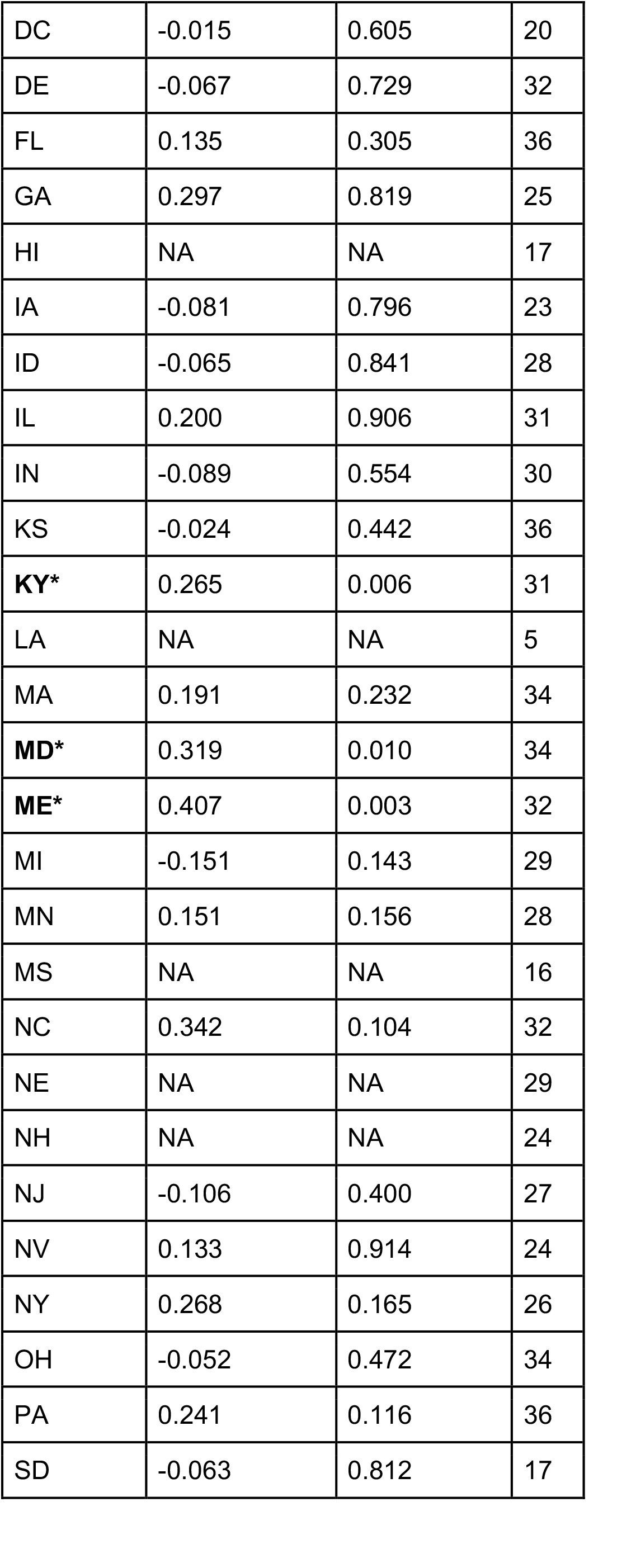
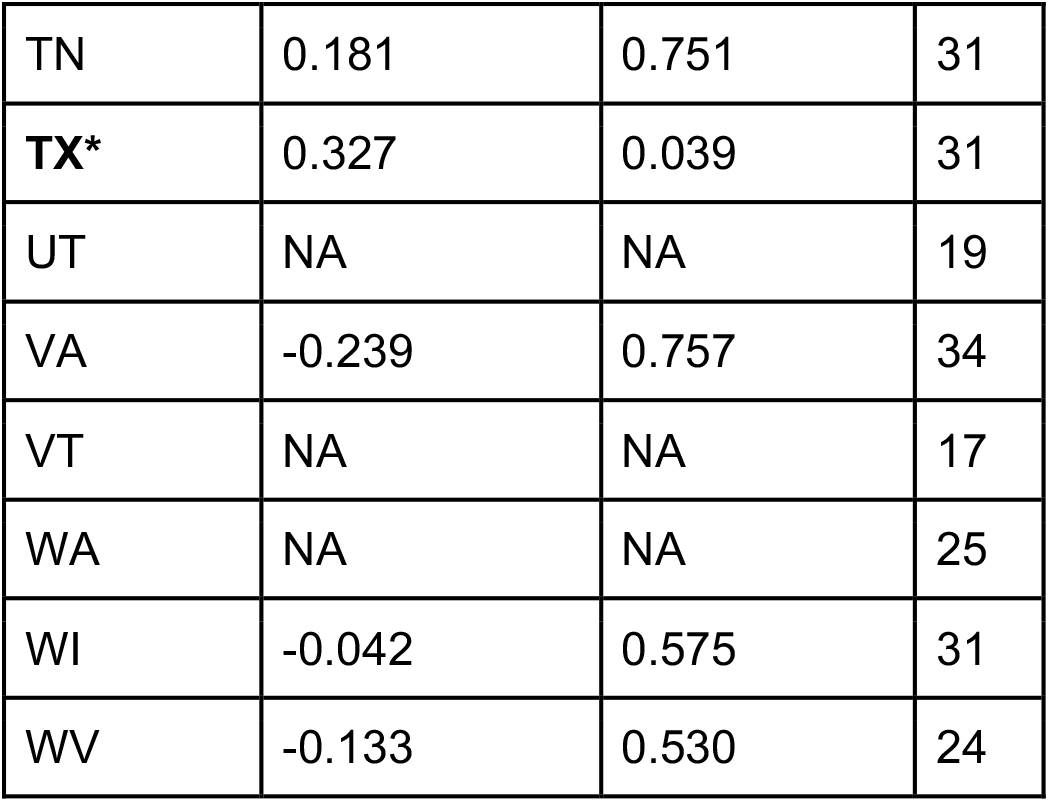
Table summarizing Kendall’s rank correlation, tau, between weekly cases recorded by NNDSS and weekly average wastewater concentrations. NA indicates that either the concentration values or case values were all 0 for a state, which meant no Kendall’s rank correlation coefficient could be calculated, n represents the number of observations in each group (state) and an asterisk next to the state abbreviation indicates that the correlation had a p-value less than 0.05. DC is Washington DC.

We calculated correlations on a national level between weekly population weighted average wastewater concentrations and weekly new cases as reported by NNDSS (Kendall’s τ = 0.20, p=0.04963). We ran the same analysis using the percentage of positive wastewater detections (Kendall’s τ = 0.33, p=0.001429,). Lastly, we measured the percentage of positive wastewater samples across the entire study for each state and compared these to the total number of cases per 100,000 people in each state during the study period (Kendall’s τ = 0.38, p=0.006683). The results are summarized in Figure 2.

**Figure 1:**
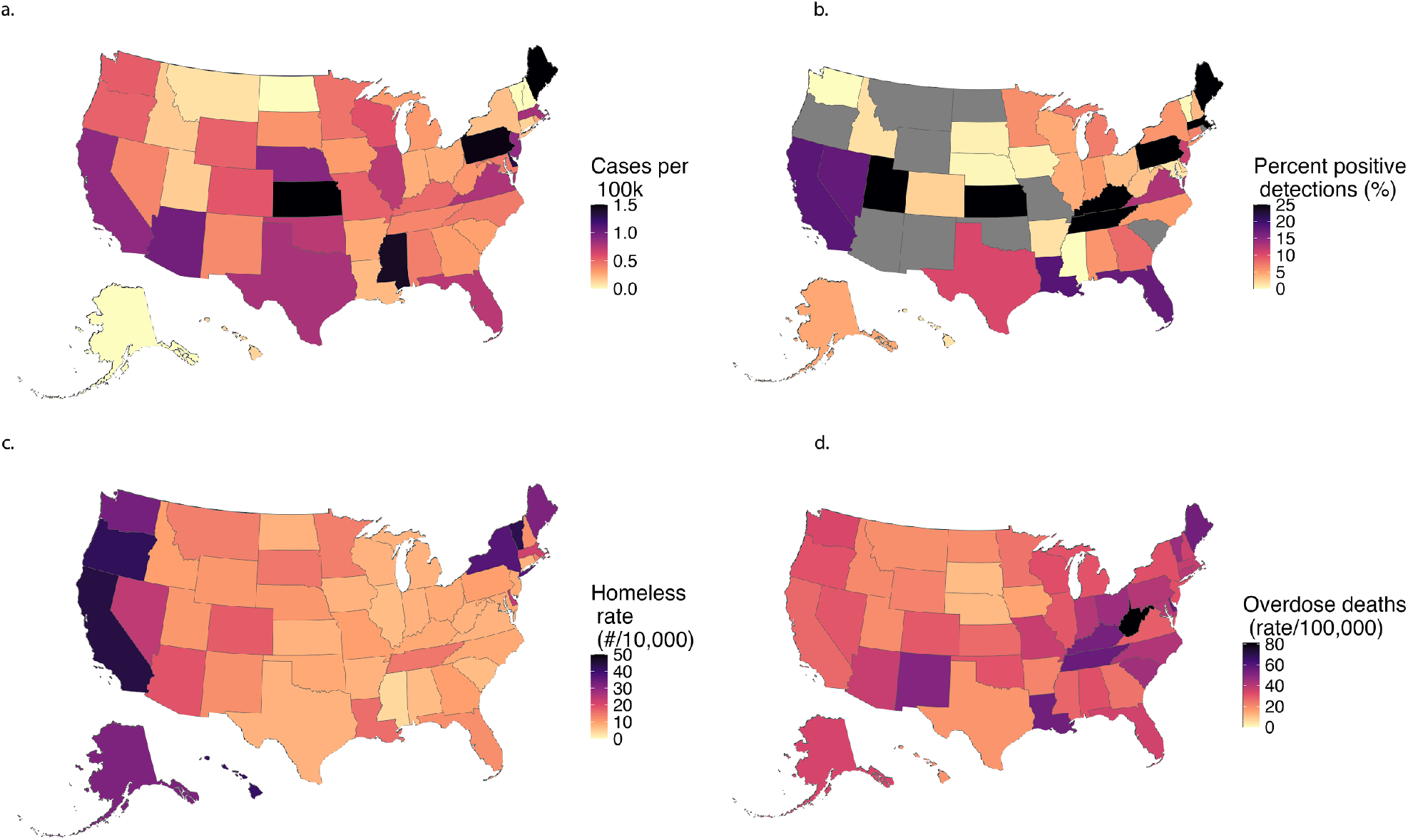
Heatmaps representing nationwide measurements used herein by state. Figure 1a represents the number of cases per million population as reported by NNDSS. Figure 1b represents the percentage of wastewater samples positive for HAV aggregated by state. Figure 1c shows the rates of homelessness across states normalized per 10,000 population, and figure 1d represents deaths attributed to drug overdoses in the CDC WONDER database normalized per 100,000 population.

**Figure 2:**
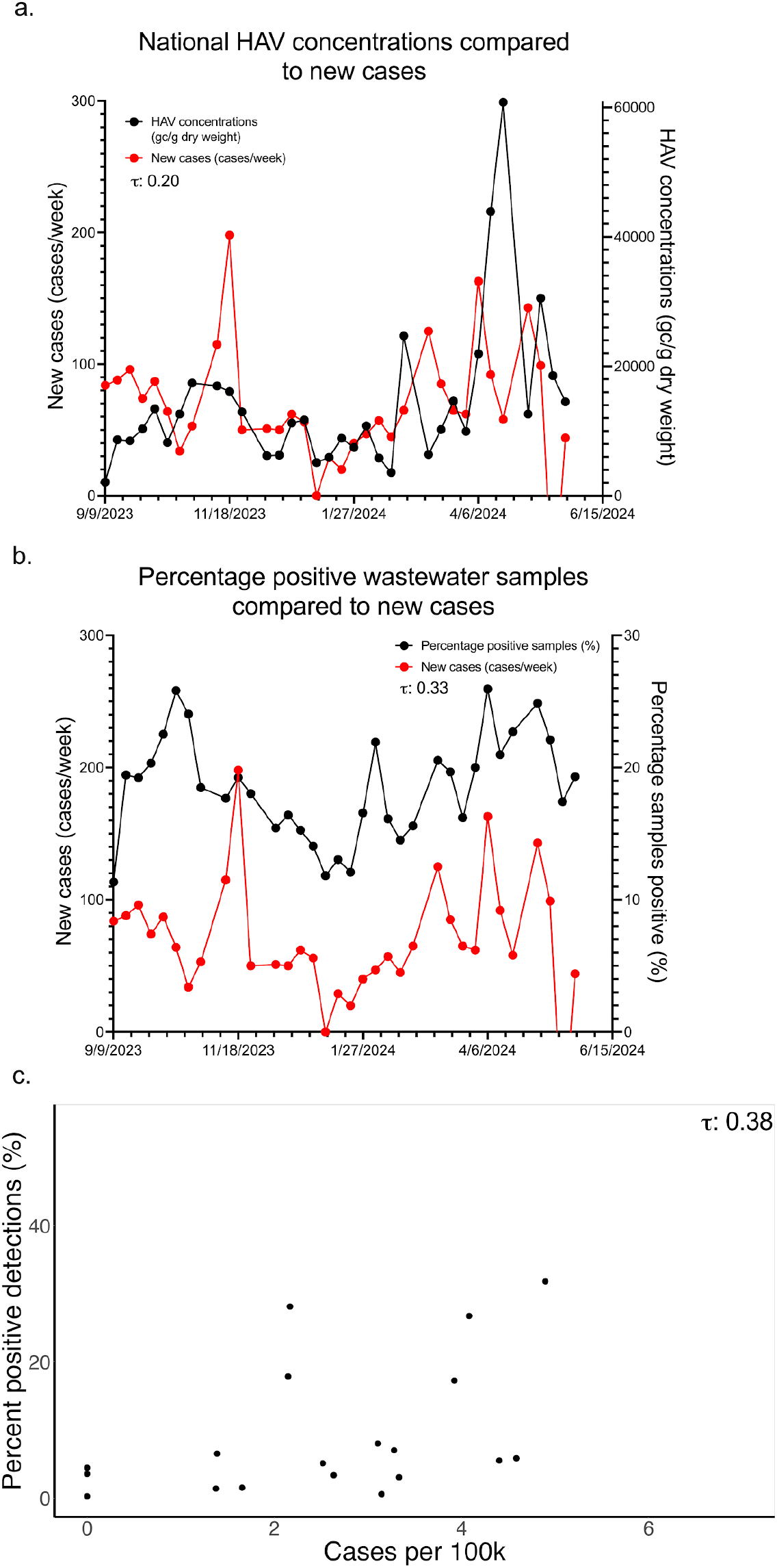
Summary of national concentrations and percent positive detections of HAV in wastewater compared to case counts nationally and cases per 100,000 for states. Figure 2a shows national, population weighted weekly HAV concentrations compared to weekly case counts as reported to NNDSS. Figure 2b compares the percentage of samples positive for HAV across all samples to case counts. Figure 2c plots a summary of detection in each state, comparing the percentage of all samples positive for HAV in the state to cases adjusted by population.

### Risk factors

We aggregated quantitative data on two common risk factors for HAV, homelessness and drug overdose deaths, to determine whether these were associated with HAV detection in wastewater. The yearly average rate of drug overdose deaths across the United States is 32.6 per 100,000 population. We then split our observations at the state level based on this, grouping all observations in areas with equal to or above average rates of drug overdoses together. This resulted in 4,169 observations in the equal to or above average overdose group and 14,836 observations in the below average overdose group. The average percentage of positive wastewater detections in the equal to or above average overdose group was 16.47% (95% CI: 15.35 - 17.60) compared to 14.4% (95% CI: 13.85 - 14.97) in the below average overdose group. These values were significantly different, with a p-value less than 0.0001. The average rate of homelessness in the US is 18 per 10,000 population. We split our data in the same way, resulting in 10,261 observations in the equal to or above average homelessness group and 11,286 in the below average homelessness group. The average percentage of positive detections of HAV in the equal to or above average homelessness group was 17.00% (95% CI: 16.28 - 17.74) while it was 11.49% (95% CI: 10.92 - 12.07) in the below average homelessness group. These values were significantly different, with a p-value less than 0.0001. These differences represent a 48% higher chance of HAV detection in wastewater in areas with high rates of homelessness and a 14% higher chance of HAV detection in areas with high rates of drug overdoses. Figure 3 summarizes these results.

**Figure 3:**
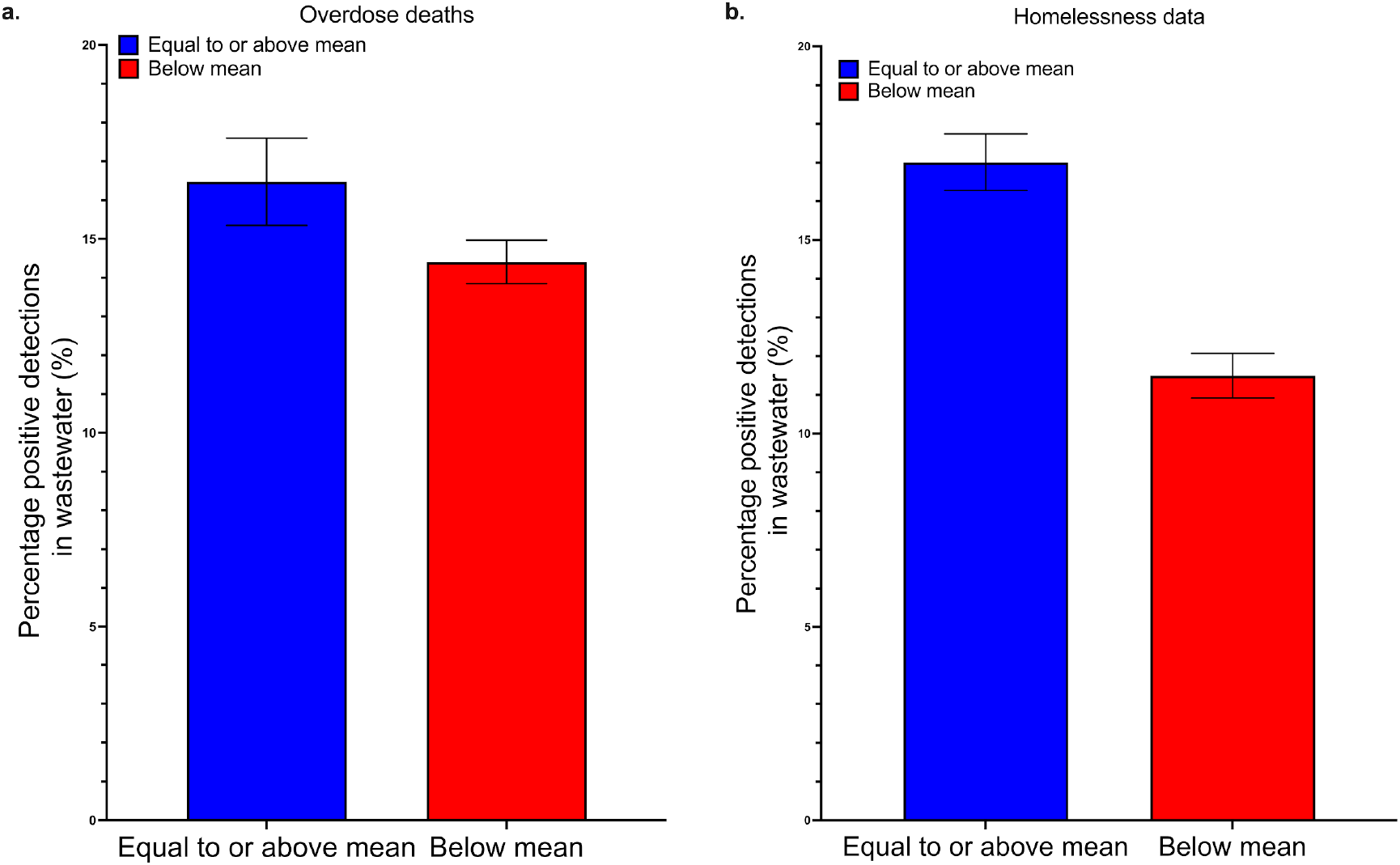
Association between risk factors on wastewater detections of HAV nucleic acids. **Figure 3a** demonstrates significantly higher wastewater detections in states with higher rates of overdose deaths. **Figure 3b** shows the association of homelessness on wastewater detections of HAV, with significantly higher positive rates in states with equal to or above median rates of unhoused individuals. Error bars represent 95% confidence intervals. Null distribution graphs are provided in the SI, Figure S3.

### Case study: Portland, Maine

Case data available through NNDSS can be limited, so we identified an area of high HAV occurrence, the state of Maine, and requested county level, daily case data from the Division of Disease Surveillance in the Maine Department of Health Human Services (MDPH). We performed detailed analytical comparisons as a case study. In September of 2023, HAV cases rose in the city of Portland in Cumberland County. In the 8 months between 1 January 2023 to 30 August 2023, a total of 3 cases were recorded. Then, in September 2023, 5 cases were identified (Figure 4). An outbreak was declared by the MDPH in October, 2023. Several exposure events were identified by MDPH, all in Cumberland County or the adjacent Androscoggin County. Three WWTP participating in our study are located in these counties, with two in Cumberland and one in Androscoggin (Portland Water District (East End Wastewater Treatment Facility), Brunswick Sewer District and Lewiston Auburn Water Pollution Control Authority). As shown in Figure 4, cases in the counties had a high degree of correlation to wastewater concentrations of HAV averaged on a monthly basis at WWTPs serving the counties. Specifically, we found a Kendall rank correlation coefficient of 0.93 and 0.97 when comparing average monthly wastewater HAV RNA concentrations in each county to monthly cases for Androscoggin and Cumberland county, respectively (p<0.001). Using weekly concentrations and weekly case numbers for these 2 counties, Kendall rank correlation coefficients were 0.53 (Androscoggin, p<0.001) and 0.42 (Cumberland, p<0.001). In Cumberland county, where wastewater monitoring began the outbreak, cross-correlation analysis demonstrated that wastewater data lead clinical surveillance data by a week (Figure 5).

**Figure 4:**
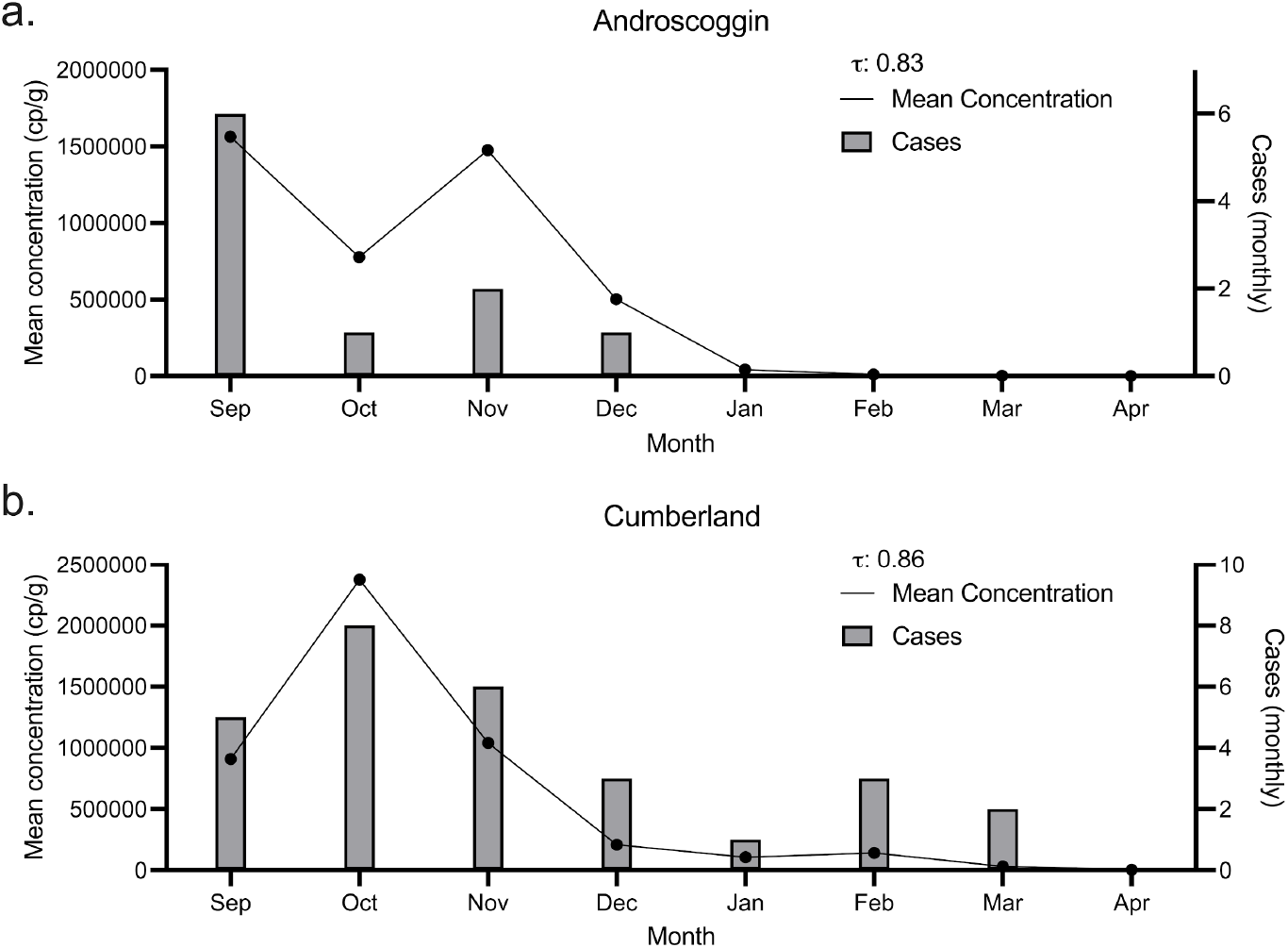
Figure demonstrating association between HAV wastewater concentrations and cases in Maine. The figures show monthly mean concentrations as a black line, and monthly cases as grey bars. Kendall’s tau is provided in the top right. Figure 4a shows these results for Androscoggin county, while figure 4b shows these results for Cumberland county.

**Figure 5:**
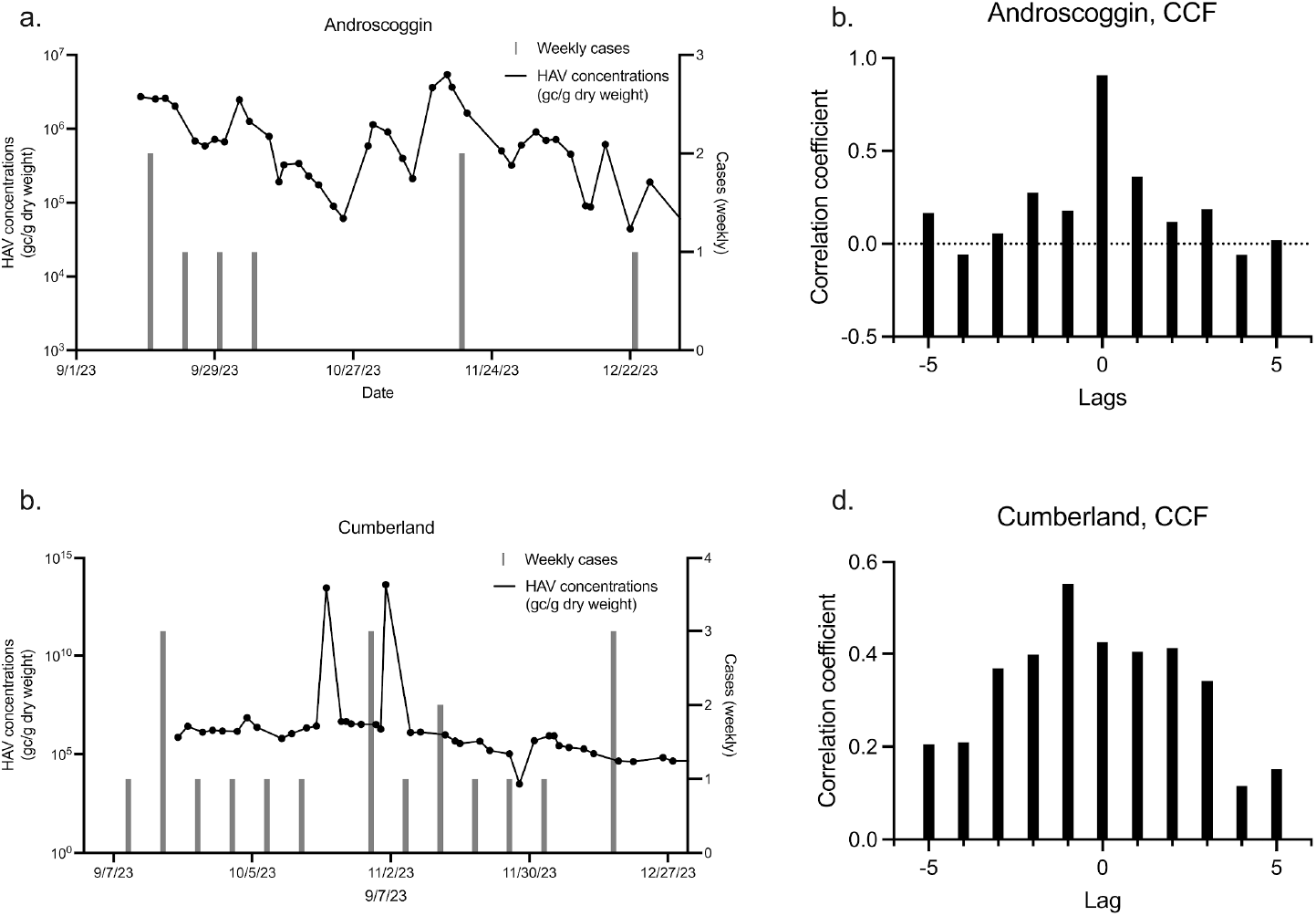
Raw wastewater concentrations compared to weekly cases, and the accompanying cross-correlation analysis. Concentrations are presented on a log10 scaled y-axis. Figures 5a and 5b are the results for Androscoggin county. Figures 5c and 5d show these results for Cumberland county.

## Discussion

Previous studies of HAV in wastewater have shown that detection of the virus in wastewater streams is feasible but have largely focused on analyses of the occupational exposure risk that these viruses represent.^39–42^ More recent studies, spurred on by the success of SARS-CoV-2 wastewater monitoring and recent HAV outbreaks in the United States, have begun studying the relationship between wastewater concentrations of HAV and laboratory-confirmed clinical cases.^14,20,43,44^ These have shown limited but promising results regarding the forecasting ability of measuring HAV in wastewater.^14,43^ These studies were limited to small spatial scales such as within a single city.^14,43,45^ We present the results of a nationwide wastewater monitoring program covering 40 states and the District of Columbia and demonstrate that HAV RNA wastewater concentrations are associated with disease occurrence in the population at the national level, within some states, and at the county level in Maine. This is despite the fact that the incident case data may be biased to cases of severe disease and may not include undiagnosed and subclinical cases. Further, we show that wastewater detection of HAV RNA is associated with socio-economic indicators of vulnerability that highlight the ability of wastewater monitoring to serve vulnerable populations, and open up the idea of using wastewater monitoring to track the effects of interventions in these populations. Lastly, the detailed case study of HAV outbreaks in Maine demonstrates a potential lead time of wastewater over public health surveillance data in one of two sites.

There were states for which HAV in wastewater solids was not significantly associated with case rates. Wastewater and case data capture different populations of individuals. Whereas wastewater captures contributions from all infected individuals, including those who may be asymptomatic, case rates only reflect severe cases that are diagnosed and reported to NNDSS; case reporting fidelity and frequency may vary by state. These differences undoubtedly affect the association between wastewater and case rates seen in many of the states.

Nationally, 13.76% of all wastewater samples were positive for HAV during the study. These results are in agreement with previous studies regarding the presence of HAV in wastewater but those other studies measured HAV in the liquid rather than solid fraction of wastewater. Fantilli et al. measured HAV concentrations in wastewater influent at a treatment plant in Cordoba, Argentina and found similar levels of variability in percentage of detections across locations and time periods (2.9% - 56.5%).^14^ They also found that HAV genotypes detected in wastewater samples matched those identified in clinical samples, further confirming that HAV in wastewater can reflect clinical infections. A similar study by Hellmer et al. suggested that due to high levels of shedding of the virus by infected patients, even a single infection could cause large spikes in wastewater concentrations followed by non-detectable concentrations.^45^ This is corroborated by our case study in Maine, where even one detected case in a month appears to reflect higher HAV concentrations. Though limited data is available on HAV in wastewater solids, Yin et al. identified one study that indicated HAV partitioning to solids.^46^

There are limited data on shedding of HAV in excretions of infected individuals, however, it has been documented in feces, sputum, and urine of infected individuals.^15–17,47^ HAV can be detected in stool up to 45 days after infection, though the concentration of these detections drops off rapidly the week after infection.^47,48^ It has been noted that in some immunocompromised patients, replication of the HAV virus can occur despite vaccination.^49^ Hundekar et al. showed that infected asymptomatic persons may shed the virus at elevated concentrations for longer periods of time than symptomatic persons, even if HAV antibodies are present.^47^ Further studies on shedding in unvaccinated and vaccinated individuals, in various bodily excretions,is necessary to further link HAV wastewater concentrations to occurrence of infections in the contributing population.^49^

The early detection of viral occurrence in a community is a key measure that allows public health officials to enact appropriate responses such as vaccination campaigns. Despite the availability of a vaccine and increasing water, sanitation, hygiene standards, HAV remains one of the most common enteric viruses in the world and is responsible for an estimated 39,000 deaths worldwide.^7,8^ Further, increasing levels of hygiene and immunity gaps in adult populations in countries such as the United States have the potential to increase the intensity of outbreaks.^3,5,12,13,50^ Wastewater monitoring represents a rapid method for the detection of infections of HAV, and the results presented in this study support the conclusion that detection in wastewater can be representative of underlying infections and case rates. Therefore, wastewater HAV detections may be useful for health professionals for making informed decisions about where to target limited resources, such as vaccination clinics. The congruence between wastewater and surveillance data, despite the latter’s limited availability and resolution, highlights an advantage of using wastewater monitoring for illnesses with limited clinical testing.

Hepatitis A is a disease closely associated with socioeconomic indicators of vulnerability.^1,12^ Worldwide, it is largely driven by lacking sanitary conditions and access to clean water.^5,8^ In the US, it is associated with two major risk factors: homelessness and drug use.^1,2,10^ To identify whether wastewater trends reflected this, drug overdose data and rates of homelessness were compiled for comparison. We then demonstrated that wastewater concentrations reflect this epidemiological trend, with significantly higher concentrations of wastewater HAV in states with higher drug overdose deaths and rates of individuals experiencing homelessness. In areas with high levels of homelessness, we were 48% more likely to detect HAV, while in areas with high rates of drug overdoses, we were 14% more likely to detect HAV. These results show that wastewater concentrations may reflect the underlying presence of HAV in a population, and that wastewater results can help identify at-risk populations, healthcare inequities, and risk factors that public health surveillance case data may miss.

To corroborate these findings, we present an in-depth analysis of wastewater concentrations and public health surveillance case data in the state of Maine during our monitoring period. Public health surveillance case data were provided at the county level by the Maine Department of Health Human Services. The results of this analysis demonstrate a high degree of agreement between signals (rank correlation coefficient of 0.93 and 0.97 in affected counties), but more importantly, identify a 1 week lead time of wastewater concentrations over public health surveillance case data, and identify sharp increases in concentrations the week before exposure events in Cumberland county. Identifying these spikes in wastewater prior to potential exposure events would allow local health officials to deploy the appropriate resources to track a point source.

Overall, we demonstrated that wastewater concentrations and percent positive detections of HAV RNA are significantly correlated with NNDSS cases nationally, in some states, and at the county level in Maine, are significantly associated with socioeconomic indicators of vulnerability, and may provide up to a week lead time over transmission events and cases. This opens up the possibility of using HAV wastewater monitoring to enact timely interventions that prevent further spread of illness, such as vaccination clinics or non-pharmaceutical interventions. These interventions are key in the prevention of future outbreaks such as the 2016 HAV outbreak in the US, and can further be used in low- and middle-income regions to identify and target the most at-risk populations for preventative measures.^3,12^ In these regions where HAV is endemic, it will also provide a way of measuring the impact of interventions such as hygiene awareness campaigns in areas with little to no clinical testing.

The work presented in this study is subject to some limitations. Public health surveillance data were limited to reported HAV cases and were likely not inclusive of all incident cases. Both the wastewater sampling and public health surveillance data are not uniform across the US or individual states, which could have introduced biases into our analyses. Case data is biased towards severe cases where individuals seek care, which underestimates disease prevalence for illnesses such as HAV, where up to 70% of cases are asymptomatic.^11,13^ By comparison, wastewater captures the entire population contributing within the sewershed even those with mild and asymptomatic cases, which is a separate and complementary measurement to public health surveillance cases. Such differences between clinical surveillance and wastewater monitoring data suggest that the two metrics might not be directly comparable, although they are complementary. Within the socioeconomic data available, we were limited to data provided at the state level versus the sewershed level, and homelessness and rates of drug overdoses vary considerably within a state. The exact magnitude of impact of these socioeconomic factors will require further investigation at smaller geographical scales.

## Data Availability

All data produced are available online at the Stanford Digital Repository (https://purl.stanford.edu/qf850cv6453).

https://purl.stanford.edu/qf850cv6453

## Acknowledgments

We thank the participating wastewater treatment plants for their samples for the project.

