## Supporting Material for "Detection of *Hepatovirus A* (HAV) in wastewater indicates widespread national distribution and association with socioeconomic indicators of vulnerability"

**Equation for population weighted HAV concentrations**

$HAV\_pop\_w = (\sum_{i=1}^{X} pop_i * HAV_{mmwr}) / (\sum_{i=1}^{X} pop_i)$ (Eq 1)

Where HAV_pop_w is the state-aggregated population weighted average of HAV concentrations. $Pop_i$ is the population served by WWTP i of X total in the state, and $HAV_{mmwr}$ is the weekly mean of HAV gene copies per gram as grouped by Morbidity and Mortality Weekly Report (MMWR) weeks.

**Multiplexing interference assessment.** We sought to confirm that the multiplexing seven assays did not interfere with HAV target quantification. We tested whether the quantification of HAV in the presence of and absence of similar concentrations of the six other targets, including those for which results are not reported in this study (influenza A and B virus, the N gene of SARS-CoV-2, respiratory syncytial virus, norovirus GII, and rotavirus), was substantially different. To do so, we first quantified three decimal dilutions of a HAV nucleic acid in the absence of any other targets using the (RT)-PCR chemistry that included primers and probes for all eight targets. Then, we quantified the same decimal dilutions of the single target in the presence of 10-100 copies per reaction of the seven other targets. Each dilution was run in a single well and no template, negative controls were included on each PCR plate. Assays were run and thresholded as described in the main paper. Results were expressed as copies per reaction and the standard deviation, as output by the instrument, were included. Results are provided in Figure S1 which suggested no interference.

**Additional details related to EMMI guidelines.** Two-hundred six (206) wastewater samples from the study were selected at random for this analysis; this represents ~1% of the samples processed in the study. The randomly selected samples were from the samples for which 10 replicate wells were run for HAV as these raw data were relatively more easily accessible. The average (standard deviation) number of partitions (droplets) for the across 10 replicate wells was 175,792 (30,304) for the reaction for HAV. The volume of the partitions, as reported by the machine vendor is 0.00085 µL. The average (standard deviation) of copies per partition for the HAV target is $4.92 \times 10^{-5}$ ($1.58 \times 10^{-4}$). An example fluorescent plot from the QX600 (6 color reader) is included in the Stanford Digital Repository with the deposited wastewater data (https://purl.stanford.edu/qf850cv6453).

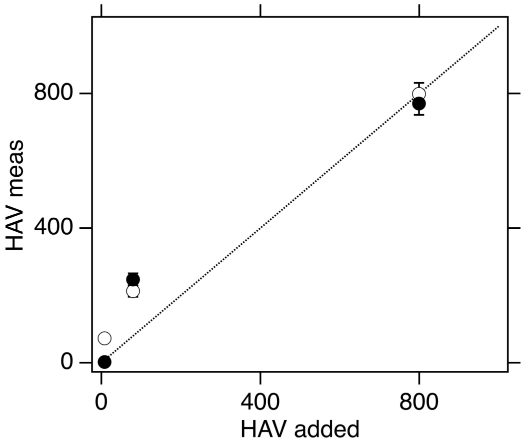


Figure S1. Multiplex assay performance. Concentrations of the HAV target in units of copies per reaction input (“HAV added”) and measured (“HAV meas”) is provided. The white symbols are for reactions without the 6 background nucleic-acid targets and black symbols include high concentrations of the 6 other nucleic-acid targets. Error bars are standard deviations, if error bars cannot be seen, then they are smaller than the symbol. The line represents the 1:1 line. NTC wells returned 0 copies of target. The markers near 0 are approximately 10 copies added.

| Study Description | Environmental Sampling | Sample Treatment | Sample Reduction | Nucleic-acid Extraction | Reverse Transcription | PCR Amplification | Analysis |
|---|---|---|---|---|---|---|---|
| Study name: HAV nation<br>Date: July 2024<br>Completed by: A. Boehm | Notes: Decribed in methods section | Notes: No sample treatment performed | Notes: Centrifugation was used, as described in the methods | Notes: Methods provided in paper. | Notes: Performed one-step RT-PCR | Notes: droplet digital PCR used | Notes: Provided in methods |

| Control Checklist | Environmental Sampling | Sample Treatment | Sample Reduction | Nucleic-acid Extraction | Reverse Transcription | PCR Amplification | |
|---|---|---|---|---|---|---|---|
| Step performed | ☑ | ☐ | ☑ | ☑ | ☑ | ☑ | |
| Step has control info | ☐ | ☐ | ☐ | ☑ | ☑ | ☑ | Negative controls |
| # of control replicates | 0 | na | 0 | 2 | 2 | 2 | |
| Control result reported | ☐ | ☐ | ☐ | ☑ | ☑ | ☑ | |
| Method for handling failed controls described | ☐ | ☐ | ☑ | ☑ | ☑ | ☑ | |
| Step has control info | ☑ | ☐ | ☑ | ☐ | ☐ | ☐ | Positive controls |
| Control identity described | ☑ | ☐ | ☑ | ☑ | ☑ | ☑ | |
| Control quantification method described | ☑ | ☐ | ☑ | ☑ | ☑ | ☑ | |
| # control replicates | internal control | na | 6 to 10 | 6 to 10 | 6 to 10 | 6 to 10 | |
| Control result reported | ☑ | ☐ | ☑ | ☑ | ☑ | ☑ | |
| Method for handling failed controls described | ☑ | ☐ | ☑ | ☑ | ☑ | ☑ | |

**Process checklist**

| Environmental Sampling | |
|---|---|
| Sample procedure | ☑ |
| Number of samples | ☑ |
| Sample amount, mean, range | ☑ |
| Sampling locations, dates, times | ☑ |
| Sample storage conditions | ☑ |

| Sample Treatment | |
|---|---|
| Treatment procedure | ☐ |
| Reagents | ☐ |

| Sample Reduction | |
|---|---|
| Reduction procedure | ☑ |
| Reagents | ☑ |
| Concentration factor | ☑ |

| Nucleic-acid Extraction | |
|---|---|
| Extraction procedure | ☑ |
| Volume or mass extracted, volume or mass obtained | ☑ |
| Extract storage conditions | ☑ |

| Reverse Transcription | |
|---|---|
| One- or two-step | ☑ |
| cDNA storage conditions (if 2 step) | ☐ |
| Reaction temperatures and times | ☑ |
| Reaction reagents and concentrations | ☑ |
| Priming method | ☑ |
| Reaction volume, added template amount | ☑ |
| RT efficiency assessment procedure (if 2-step) | ☐ |
| RT control description (if two-step) | ☐ |
| RT efficiency reported (if 2-step) | ☐ |

| qPCR or dPCR | |
|---|---|
| Target gene name, amplicon length | ☑ |
| Thermocycling temp and times | ☑ |
| Master mix composition: vendors, concentrations | ☑ |
| Additives: vendors, composition | ☑ |
| Template amount added, pre-treatment (if any) | ☑ |
| Primers: sequences, concentrations, vendors, references | ☑ |
| Amplicon confirmation method (probe, melt curve details, etc) | ☑ |
| Probe sequence, concentration, vendor, reference | ☑ |
| Instrumentation | ☑ |
| Inhibition assessment procedure | ☑ |
| Inhibition control description (if used) | ☐ |
| Number of samples tested and found inhibited | ☐ |
| Equivalent volume of sample analyzed | ☑ |

| Analysis- dPCR | |
|---|---|
| Threshold settings | ☑ |
| Technical replicates, number, well merging | ☑ |
| Partitions measured, number, mean, variance | ☑ |
| Partition volume | ☑ |
| Target copies per partition, mean, variance | ☑ |
| Program used for dPCR analysis | ☑ |
| Explanation of control results, example plots | ☑ |

| Analysis- qPCR | |
|---|---|
| Technical replicates, number, calculations | ☐ |
| Calibration standards, description, source | ☐ |
| Method of quantifying standards | ☐ |
| Calibration curve slope | ☐ |
| Calibration curve R2 | ☐ |
| Lowest standard measured or 95% LOD | ☐ |
| Cq value determination methods | ☐ |

Note to users: This checklist is provided as guidance for best practices for reporting, but is not meant to be prescriptive. Not all items in the check list will apply to all studies. Please see Borchardt et al. The Environmental

Version 3.0

This version maintained by Borchardt, Boehm, Salit, Noble, Wiggington, Spencer

**Date: 18 October 2023**

Figure S2. EMMI guidelines[1] checklist.

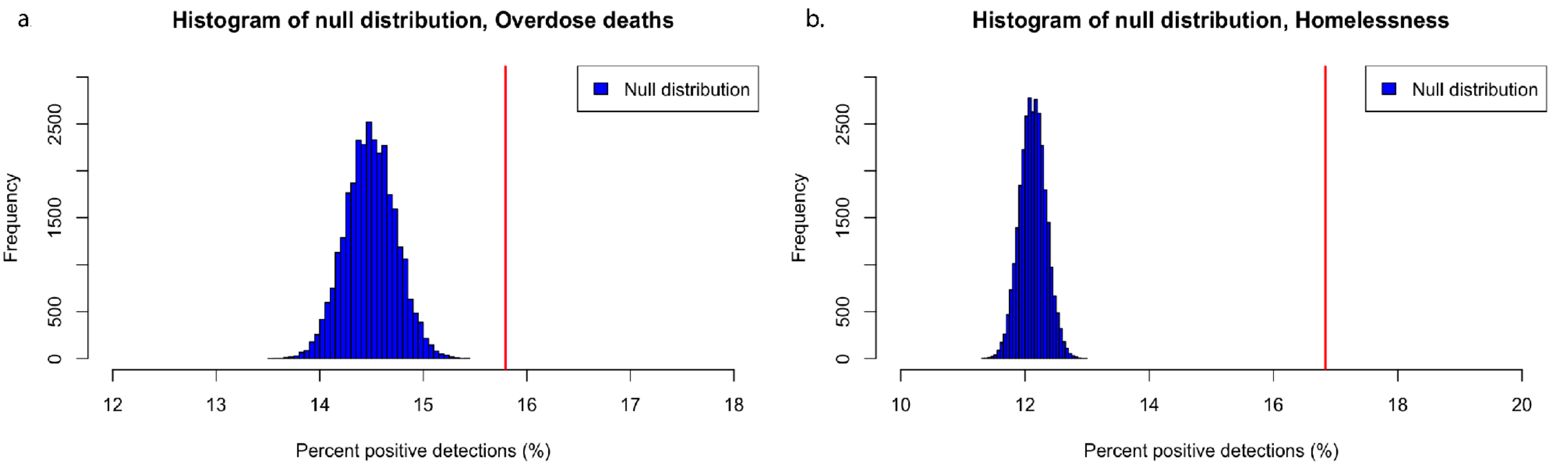


Figure S3. Null distributions compared to test statistics. Figure S3A shows the null distribution in blue bars for percent positive detections of HAV. The red line represents our test statistic. In both analyses, the test statistic is far outside the null distribution.

Table S1. Plant characteristics including sample type and population served. Plants marked with an asterisk (*) are plants where 10 replicate wells were used for digital droplet PCR, the remaining plants had 6 wells used to measure HAV, and 2 for PMMoV and BCoV. The site name, location (town, and state), state where plant is located, and whether the sample originally supplied to us was a 24 hour composite sample of liquid influent ("liquid") or a sample from the primary clarifier ("solids").

| Site Name | Location | State | Sample Type | Population served |
|---|---|---|---|---|
| Valley Creek Water Reclamation Facility | Bessemer, AL | Alabama | Liquids | 225,000 |
| Cahaba River Water Reclamation Facility | Cahaba River, Birmingham, AL | Alabama | Liquids | 95,000 |
| Five Mile Creek Water Reclamation Facility | Fultondale, AL | Alabama | Liquids | 77,000 |
| Turkey Creek Water Reclamation Facility | Pinson, AL | Alabama | Liquids | 30,000 |
| Village Creek Water Reclamation Facility | Village Creek, Birmingham, AL | Alabama | Liquids | 200,000 |
| John M. Asplund Water Pollution Control Facility | Anchorage, AK | Alaska | Liquids | 220,000 |
| City of Harrison Wastewater Treatment Plant | Harrison, AR | Arkansas | Liquids | 15,000 |
| Coastal Treatment Plant | Coastal, Laguna Niguel, CA | California | Liquids | 48,000 |
| Central Contra Costa Sanitary District | Contra Costa County, CA | California | Solids | 484,800 |
| City of Davis Wastewater Treatment Plant | Davis, CA | California | Solids | 68,000 |
| Esparto Wastewater Treatment Facility | Esparto, CA | California | Liquids | 4,006 |
| Fairfield-Suisun Sewer District | Fairfield, CA | California | Solids | 155,000 |
| [Fremont Basin] - Raymond A. Boege Alvarado WWTP | Fremont, CA | California | Liquids | 229,476 |
| South County Regional Wastewater Authority * | Gilroy, CA | California | Solids | 110,338 |
| Sewer Authority Mid-Coastside | Half Moon Bay, CA | California | Liquids | 28,000 |

| | | | | |
|---|---|---|---|---|
| City of Hollister Domestic Water Recycling Facility | Hollister, CA | California | Liquids | 42,000 |
| Valley Sanitary District | Indio, CA | California | Solids | 91,765 |
| JB Latham Treatment Plant | JB Latham, Laguna Niguel, CA | California | Liquids | 120,000 |
| Lancaster Water Reclamation Plant | Lancaster, CA | California | Liquids | 200,000 |
| Las Gallinas Valley Sanitary District | Las Gallinas, San Rafael, CA | California | Liquids | 30,000 |
| Lompoc Regional Wastewater Reclamation Plant | Lompoc, CA | California | Liquids | 69,290 |
| Joint Water Pollution Control Plant | Los Angeles County, CA | California | Liquids | 3,500,000 |
| Hyperion Water Reclamation Plant (HWRP) | Los Angeles, CA | California | Liquids | 4,000,000 |
| Los Banos Wastewater Treatment Plant | Los Banos, CA | California | Liquids | 42,000 |
| City of Madera, Wastewater Treatment Plant | Madera, CA | California | Solids | 67,944 |
| Mammoth Community Water District | Mammoth, CA | California | Liquids | 35,000 |
| Monterey One Water - Regional Treatment Plant | Marina, CA | California | Liquids | 262,000 |
| Merced Wastewater Treatment Plant | Merced, CA | California | Solids | 91,000 |
| Sewerage Agency of Southern Marin Wastewater Treatment Plant | Mill Valley, CA | California | Solids | 30,000 |
| Modesto's Sutter Primary Treatment Facility | Modesto, CA | California | Solids | 230,000 |
| Soscol Water Recycling Facility | Napa, CA | California | Solids | 83,300 |
| [Newark Basin] - Raymond A. Boege Alvarado WWTP | Newark, CA | California | Liquids | 47,229 |
| Novato Sanitary District | Novato, CA | California | Liquids | 53,000 |
| East Bay Municipal Utility District | Oakland, CA | California | Solids | 740,000 |
| Oceanside Water Pollution Control Plant * | Oceanside, San Francisco, CA | California | Solids | 250,000 |

| | | | | |
|---|---|---|---|---|
| Regional Water Recycling Plant No.1 (RP-1) | Ontario, CA | California | Solids | 890,000 |
| Calera Creek Water Recycling Plant | Pacifica, CA | California | Liquids | 40,000 |
| Palo Alto Regional Water Quality Control Plant * | Palo Alto, CA | California | Solids | 236,000 |
| City of Paso Robles Wastewater Treatment Plant | Paso Robles, CA | California | Solids | 31,037 |
| Ellis Creek Water Recycling Facility | Petaluma, CA | California | Liquids | 65,000 |
| Silicon Valley Clean Water * | Redwood City, CA | California | Solids | 199,000 |
| Regional Treatment Plant | Regional, Laguna Niguel, CA | California | Liquids | 129,000 |
| Riverside Water Quality Control Plant | Riverside, CA | California | Liquids | 350,000 |
| Sacramento Regional Wastewater Treatment Plant * | Sacramento, CA | California | Solids | 1,480,000 |
| E.W. Blom Point Loma Wastewater Treatment Plant | San Diego, CA | California | Liquids | 2,200,000 |
| San Jose-Santa Clara Regional Wastewater Facility * | San Jose, CA | California | Solids | 1,500,000 |
| City of San Leandro Water Pollution Control Plant | San Leandro, CA | California | Liquids | 50,000 |
| City of San Mateo & Estero M.I.D. Water Quality Control Plant | San Mateo, CA | California | Solids | 150,000 |
| Central Marin Sanitation Agency | San Rafael, CA | California | Liquids | 104,250 |
| City of Santa Cruz WTF - County Influent | Santa Cruz County, CA | California | Solids | 160,000 |
| City of Santa Cruz WTF - City Influent | Santa Cruz, CA | California | Solids | 160,000 |
| City of Santa Rosa, Laguna Treatment Plant | Santa Rosa, CA | California | Solids | 230,000 |
| Sausalito-Marin City Sanitary District | Sausalito, CA | California | Liquids | 18,000 |
| South Bay International Wastewater Treatment Plant | South San Diego, CA | California | Solids | 1,600,000 |
| Southeast Waste Pollution Control Plant * | Southeast San Francisco, CA | California | Solids | 750,000 |

| | | | | |
|---|---|---|---|---|
| City of Sunnyvale Water Pollution Control Plant * | Sunnyvale, CA | California | Solids | 153,000 |
| Turlock Regional Water Quality Control Facility | Turlock, CA | California | Liquids | 86,000 |
| [Union City Basin] - Raymond A. Boege Alvarado WWTP | Union City, CA | California | Liquids | 68,150 |
| Vallejo Flood and Wastewater District Wastewater Treatment Plant | Vallejo, CA | California | Liquids | 121,000 |
| West County Wastewater District | West Contra Costa County, CA | California | Liquids | 100,000 |
| Central Marin Sanitation Agency - West Railroad | West Railroad, San Rafael, CA | California | Liquids | 25,000 |
| Windsor Wastewater Treatment, Reclamation, and Disposal Facility | Windsor, CA | California | Liquids | 28,000 |
| Winters - East Street Pump Station | Winters, CA | California | Liquids | 7,286 |
| Woodland Water Pollution Control Facility | Woodland, CA | California | Liquids | 59,000 |
| Parker Water and Sanitation District North Water Reclamation Facility | North, Parker, CO | Colorado | Liquids | 35,000 |
| Parker Water and Sanitation District South Water Reclamation Facility | South, Parker, CO | Colorado | Liquids | 25,000 |
| Stamford Water Pollution Control Authority (WPCA) | Stamford, CT | Connectic ut | Solids | 140,000 |
| Seaford Wastewater Treatment Facility | Seaford, DE | Delaware | Solids | 13,172 |
| Altamonte Springs Regional Water Reclamation Facility | Altamonte Springs, FL | Florida | Liquids | 95,000 |
| Eastern Water Reclamation Facility | Eastern, Orange County, FL | Florida | Solids | 195,299 |
| Loxahatchee River Environmental Control District | Jupiter, FL | Florida | Liquids | 90,000 |
| MDWASD Central District WWTP | Key Biscayne, FL | Florida | Liquids | 829,725 |
| MDWASD North District WWTF | North Miami, FL | Florida | Liquids | 776,150 |
| Northeast Water Reclamation Facility | Northeast, Saint Petersburg, FL | Florida | Liquids | 89,847 |

| | | | | |
|---|---|---|---|---|
| Northwest Water Reclamation Facility | Northwest, Orange County, FL | Florida | Solids | 66,690 |
| Northwest Water Reclamation Facility | Northwest, Saint Petersburg, FL | Florida | Liquids | 94,218 |
| MDWASD South District WWTF | South Miami, FL | Florida | Liquids | 920,528 |
| South Water Reclamation Facility | South, Orange County, FL | Florida | Solids | 183,009 |
| Hamlin Water Reclamation Facility | Southwest, Orange County, FL | Florida | Solids | 50,000 |
| Southwest Water Reclamation Facility | Southwest, Saint Petersburg, FL | Florida | Liquids | 47,790 |
| TPSmith Water Reclamation Facility | Tallahassee, FL | Florida | Liquids | 212,065 |
| Big Creek Water Reclamation Facility | Big Creek, Roswell, GA | Georgia | Liquids | 189,593 |
| Camp Creek Water Reclamation Facility | College Park, GA | Georgia | Liquids | 73,821 |
| South Columbus Water Resources Facility | Columbus, GA | Georgia | Solids | 278,000 |
| Johns Creek Environmental Campus | Johns Creek, Roswell, GA | Georgia | Liquids | 84,486 |
| Little River Water Reclamation Facility | Little River, Roswell, GA | Georgia | Liquids | 12,818 |
| RM Clayton Water Reclamation Center | RM Clayton, Atlanta, GA | Georgia | Liquids | 294,660 |
| South River Water Reclamation Center | South River, Atlanta, GA | Georgia | Liquids | 105,160 |
| Utoy Creek Water Reclamation Center | Utoy Creek, Atlanta, GA | Georgia | Liquids | 70,887 |
| Hilo Wastewater Treatment Plant | Hilo, HI | Hawaii | Liquids | 16,257 |
| Honouliuli Wastewater Treatment Plant | Honouliuli, Honolulu, HI | Hawaii | Liquids | 300,000 |
| Kailua Regional Wastewater Treatment Plant | Kailua, Honolulu, HI | Hawaii | Liquids | 90,000 |
| Sand Island Wastewater Treatment Plant | Sand Island, Honolulu, HI | Hawaii | Liquids | 390,000 |
| Wahiawa Wastewater Treatment Plant | Wahiawa, Honolulu, HI | Hawaii | Liquids | 18,000 |

| | | | | |
|---|---|---|---|---|
| Waianae Wastewater Treatment Plant | Waianae, Honolulu, HI | Hawaii | Liquids | 44,000 |
| City of Coeur d'Alene Water Resource Recovery Facility | Coeur d'Alene, ID | Idaho | Solids | 50,540 |
| Lander Street Water Renewal Facility | Lander Street, Boise, ID | Idaho | Liquids | 108,556 |
| West Boise Water Renewal Facility | West Boise, ID | Idaho | Liquids | 186,901 |
| Glenbard Wastewater Authority | Glen Ellyn, IL | Illinois | Solids | 86,000 |
| Wheaton Sanitary District | Wheaton, IL | Illinois | Solids | 63,000 |
| Dillman Road WWTP | Bloomington, IN | Indiana | Liquids | 56,090 |
| City of Carmel WWTP | Carmel, IN | Indiana | Solids | 86,000 |
| Jeffersonville Downtown WWTP | Downtown, Jeffersonville, IN | Indiana | Liquids | 25,000 |
| North Water Reclamation Facility | North, Jeffersonville, IN | Indiana | Liquids | 25,000 |
| City of South Bend Wastewater Treatment Plant | South Bend, IN | Indiana | Liquids | 130,000 |
| City of Clinton | Clinton, IA | Iowa | Solids | 29,300 |
| Coralville Wastewater Treatment Facility | Coralville, IA | Iowa | Liquids | 23,000 |
| City of Marshalltown Water Pollution Control Plant | Marshalltown, IA | Iowa | Liquids | 27,400 |
| Muscatine STP | Muscatine, IA | Iowa | Solids | 24,400 |
| Ottumwa WPCF | Ottumwa, IA | Iowa | Liquids | 25,529 |
| Municipal Wastewater Treatment Plant No. 1 (Kaw Point) | Kaw Point, Kansas City, KS | Kansas | Solids | 90,000 |
| Lawrence Kansas River Wastewater Treatment Facility | Lawrence, KS | Kansas | Solids | 80,000 |
| Kansas City Treatment Plant #20 | P20, Kansas City, KS | Kansas | Solids | 35,000 |
| Salina Wastewater Treatment Plant | Salina, KS | Kansas | Solids | 47,000 |
| Wolcott Wastewater Treatment Facility | Wolcott, Kansas City, KS | Kansas | Liquids | 15,000 |
| Morris Forman Water Quality Treatment Center | Louisville, KY | Kentucky | Solids | 423,913 |

| | | | | |
|---|---|---|---|---|
| SWBNO East Bank Wastewater Treatment Plant | New Orleans | Louisiana | Liquids | 333,400 |
| SWBNO West Bank Wastewater Treatment Plant | New Orleans | Louisiana | Liquids | 50,500 |
| City of Bangor Wastewater Treatment Plant | Bangor, ME | Maine | Solids | 40,000 |
| Brunswick Sewer District | Brunswick, ME | Maine | Liquids | 10,000 |
| Lewiston Auburn Water Pollution Control Authority | Lewiston, ME | Maine | Liquids | 60,000 |
| Portland Water District (East End Wastewater Treatment Facility) | Portland, ME | Maine | Liquids | 65,000 |
| York Sewer District | York, ME | Maine | Liquids | 10,000 |
| Hagerstown Wastewater Treatment Plant | Hagerstown, MD | Maryland | Liquids | 90,000 |
| Marlay Taylor Water Reclamation Facility | Hollywood, MD | Maryland | Liquids | 55,000 |
| Deer Island Treatment Plant | Boston, MA | Massachusetts | Solids | 2,400,000 |
| Upper Blackstone Clean Water | Millbury, MA | Massachusetts | Liquids | 250,000 |
| City of Ann Arbor Wastewater Treatment Plant | Ann Arbor, MI | Michigan | Liquids | 125,000 |
| Jackson Wastewater Treatment Plant | Jackson, MI | Michigan | Solids | 90,000 |
| Grandville Clean Water Plant | Jenison, MI | Michigan | Solids | 75,000 |
| Mt. Pleasant WRRF | Mt. Pleasant, MI | Michigan | Liquids | 21,690 |
| Traverse City Regional Waste Water Treatment Plant | Traverse City, MI | Michigan | Liquids | 30,623 |
| City of Warren Wastewater Treatment Plant | Warren, MI | Michigan | Liquids | 140,000 |
| City of Mankato Water Resource Recovery Facility (WRRF) | Mankato, MN | Minnesota | Solids | 70,000 |
| Red Wing Wastewater Treatment Facility | Red Wing, MN | Minnesota | Solids | 16,000 |

| | | | | |
|---|---|---|---|---|
| City Of Rochester MN Water Reclamation Plant | Rochester, MN | Minnesota | Solids | 120,000 |
| St. Cloud Nutrient, Energy and Water Recovery Facility | St. Cloud, MN | Minnesota | Liquids | 120,000 |
| 7 C- Pascagoula Moss Point POTW (Vancleave, MS) | Jackson County, MS | Mississippi | Liquids | 34,333 |
| 2C-Gautier POTW (Vancleave, MS) | Jackson County, MS | Mississippi | Liquids | 19,008 |
| Northeast Water Resource Recovery Facility | Northeast, Lincoln, NE | Nebraska | Liquids | 60,000 |
| Theresa Street Water Resource Recovery Facility | Theresa Street, Lincoln, NE | Nebraska | Liquids | 240,000 |
| Clark County Water Reclamation District (CCWRD) Flamingo Water Resource Center (FWRC) | Las Vegas, NV | Nevada | Liquids | 990,000 |
| City of Dover Wastewater Treatment Facility | Dover, NH | New Hampshire | Liquids | 30,000 |
| Hall Street Wastewater Treatment Plant | Hall Street, Concord, NH | New Hampshire | Liquids | 45,000 |
| Penacook Wastewater Treatment Facility | Penacook, Concord, NH | New Hampshire | Liquids | 4,000 |
| South Monmouth Regional Sewerage Authority | Belmar, NJ | New Jersey | Solids | 52,672 |
| Cumberland County Utilities Authority | Bridgeton, NJ | New Jersey | Liquids | 50,000 |
| The Somerset Raritan Valley Sewerage Authority | Bridgewater, NJ | New Jersey | Liquids | 130,000 |
| Passaic Valley Sewerage Commission | Newark, NJ | New Jersey | Solids | 1,500,000 |
| Township of Ocean Sewerage Authority | Oakhurst, NJ | New Jersey | Liquids | 50,000 |
| Bayshore Regional Sewerage Authority | Union Beach, NJ | New Jersey | Solids | 100,000 |

| | | | | |
|---|---|---|---|---|
| Ithaca Area Wastewater Treatment Facility | Ithaca, NY | New York | Liquids | 90,000 |
| City of Oswego Wastewater Treatment Plant | Oswego, NY | New York | Liquids | 30,000 |
| Johnnie Mosley Regional Water Reclamation Facility | Kinston, NC | North Carolina | Liquids | 25,000 |
| City of Wilson - Hominy Creek Water Reclamation Facility | Wilson, NC | North Carolina | Solids | 50,000 |
| Archie Elledge WWTP | Winston-Salem, NC | North Carolina | Liquids | 92,000 |
| Akron Water Reclamation Facility | Akron, OH | Ohio | Solids | 365,000 |
| City of Youngstown Wastewater Treatment Plant | Youngstown, OH | Ohio | Solids | 174,000 |
| DELCORA Western Regional Treatment Plant | Chester, PA | Pennsylvania | Liquids | 220,000 |
| Capital Region Water AWTF | Harrisburg, PA | Pennsylvania | Liquids | 125,000 |
| Penn State Water Treatment Facility | Penn State University Park | Pennsylvania | Liquids | 16,000 |
| City of Yankton Wastewater Treatment Facility | Yankton, SD | South Dakota | Liquids | 20,000 |
| Moccasin Bend WWTP | Chattanooga, TN | Tennessee | Liquids | 400,000 |
| M.C. Stiles Wastewater Treatment Facility | Memphis, TN | Tennessee | Liquids | 300,000 |
| DCWT Dallas | Dallas Central, Dallas, TX | Texas | Liquids | 270,000 |
| City of Gainesville Wastewater Treatment Plant | Gainesville, TX | Texas | Liquids | 17,300 |
| City of Garland Rowlett Creek WWTP | Garland, TX | Texas | Solids | 200,000 |
| Hollywood Road WWTP | Hollywood Road, Amarillo, TX | Texas | Liquids | 60,000 |
| River Road WWTP | River Road, Amarillo, TX | Texas | Liquids | 140,000 |
| South Laredo WWTP | South, Laredo, TX | Texas | Liquids | 120,000 |

| | | | | |
|---|---|---|---|---|
| Southside Wastewater Treatment Plant (City of Dallas) | Southside, Dallas, TX | Texas | Liquids | 421,700 |
| Duck Creek Wastewater Treatment Plant | Sunnyvale, TX | Texas | Solids | 186,000 |
| DCWT White Rock | White Rock Central, Dallas, TX | Texas | Liquids | 630,000 |
| Wichita Falls Resource Recovery Facility | Wichita Falls, TX | Texas | Solids | 90,000 |
| SJRA WWTF No.1 | Woodlands SJRA WWTF No. 1, TX | Texas | Liquids | 65,000 |
| SJRA WWTF No.2 | Woodlands SJRA WWTF No. 2, TX | Texas | Liquids | 70,000 |
| SJRA WWTF No.3 | Woodlands SJRA WWTF No. 3, TX | Texas | Liquids | 15,000 |
| Zacate Creek WWTP | Zacate Creek, Laredo, TX | Texas | Liquids | 140,000 |
| Central Valley Water Reclamation Facility | Central Salt Lake Valley, UT | Utah | Solids | 600,000 |
| Provo City Water Reclamation Facility | Provo, UT | Utah | Solids | 115,000 |
| City of Essex Junction Wastewater Treatment Facility | Essex Junction, VT | Vermont | Solids | 30,000 |
| Montpelier Water Resource Recovery Facility | Montpelier, VT | Vermont | Solids | 10,100 |
| South Burlington-Airport Parkway WWTF | South Burlington, VT | Vermont | Liquids | 16,000 |
| Aquia Wastewater Treatment Facility | Aquia, Stafford, VA | Virginia | Solids | 100,000 |
| Town of Hillsville Wastewater Treatment Plant | Hillsville, VA | Virginia | Solids | 3,000 |
| Little Falls Run Wastewater Treatment Facility | Little Falls Run, Stafford, VA | Virginia | Solids | 50,000 |
| City of Snohomish WWTP | Snohomish, WA | Washington | Liquids | 10,150 |
| City of Wheeling, Water Pollution Control Division | Wheeling, WV | West Virginia | Liquids | 100,000 |

| | | | | |
|---|---|---|---|---|
| Wausau Waterworks Wastewater Treatment Facility | Wausau, WI | Wisconsin | Solids | 44,000 |
| Blue Plains Advanced Wastewater Treatment Tlant | Washington DC | District of Columbia | Liquids | 2,000,000 |

Table S2. Percentage of samples positive for HAV by plant. All samples collected during the study period were aggregated for each treatment plant. Plants are ordered by state and then alphabetically.

| **Wastewater Treatment Plant** | **State** | **Percentage of samples positive for HAV (%)** |
|---|---|---|
| John M. Asplund Water Pollution Control Facility | AK | 5.40540541 |
| Mendenhall Wastewater Treatment Plant | AK | 0 |
| Cahaba River Water Reclamation Facility | AL | 0 |
| Five Mile Creek Water Reclamation Facility | AL | 0.89285714 |
| Turkey Creek Water Reclamation Facility | AL | 3.66972477 |
| Valley Creek Water Reclamation Facility | AL | 0.87719298 |
| Village Creek Water Reclamation Facility | AL | 27.8846154 |
| City of Harrison Wastewater Treatment Plant | AR | 1.7699115 |
| [Fremont Basin] - Raymond A. Boege Alvarado WWTP | CA | 7.57575758 |
| [Newark Basin] - Raymond A. Boege Alvarado WWTP | CA | 0 |
| [Union City Basin] - Raymond A. Boege Alvarado WWTP | CA | 1.36986301 |
| A.K. Warren Water Resource Facility | CA | 60.1769912 |
| Calera Creek Water Recycling Plant | CA | 0 |
| Central Contra Costa Sanitary District | CA | 16.0377358 |
| Central Marin Sanitation Agency | CA | 0 |
| Central Marin Sanitation Agency - West Railroad | CA | 1.75438596 |

| | | |
|---|---|---|
| City of Davis Wastewater Treatment Plant | CA | 0.92592593 |
| City of Hollister Domestic Water Recycling Facility | CA | 0 |
| City of Madera, Wastewater Treatment Plant | CA | 0 |
| City of Paso Robles Wastewater Treatment Plant | CA | 5.35714286 |
| City of San Leandro Water Pollution Control Plant | CA | 4.38596491 |
| City of San Mateo & Estero M.I.D. Water Quality Control Plant | CA | 10.619469 |
| City of Santa Cruz WTF - City Influent | CA | 0.9009009 |
| City of Santa Cruz WTF - County Influent | CA | 0 |
| City of Santa Rosa, Laguna Treatment Plant | CA | 48.5981308 |
| City of Sunnyvale Water Pollution Control Plant | CA | 50.3937008 |
| Coastal Treatment Plant | CA | 0 |
| CODIGA | CA | 0 |
| E.W. Blom Point Loma Wastewater Treatment Plant | CA | 36.6071429 |
| East Bay Municipal Utility District | CA | 34.2105263 |
| Ellis Creek Water Recycling Facility | CA | 2.77777778 |
| Esparto Wastewater Treatment Facility | CA | 0 |
| Fairfield-Suisun Sewer District | CA | 20.1754386 |
| Hyperion Water Reclamation Plant (HWRP) | CA | 71.6814159 |
| JB Latham Treatment Plant | CA | 3.47826087 |

| | | |
|---|---|---|
| Lancaster Water Reclamation Plant | CA | 0 |
| Las Gallinas Valley Sanitary District | CA | 12.2807018 |
| Lompoc Regional Wastewater Reclamation Plant | CA | 6.19469027 |
| Los Banos Wastewater Treatment Plant | CA | 0 |
| Mammoth Community Water District | CA | 0 |
| Merced Wastewater Treatment Plant | CA | 6.14035088 |
| Modesto's Sutter Primary Treatment Facility | CA | 3.6036036 |
| Monterey One Water - Regional Treatment Plant | CA | 14.8514851 |
| Novato Sanitary District | CA | 1.73913043 |
| Oceanside Water Pollution Control Plant | CA | 25.3012048 |
| Palo Alto Regional Water Quality Control Plant | CA | 33.7121212 |
| Regional Treatment Plant | CA | 0 |
| Regional Water Recycling Plant No.1 (RP-1) | CA | 26.7857143 |
| Riverside Water Quality Control Plant | CA | 7.89473684 |
| Sacramento Regional Wastewater Treatment Plant | CA | 31.4393939 |
| San Jose-Santa Clara Regional Wastewater Facility | CA | 50.1886792 |
| Sausalito-Marin City Sanitary District | CA | 1.92307692 |
| Sewer Authority Mid-Coastside | CA | 14.5454545 |
| Sewerage Agency of Southern | CA | 4.42477876 |

| | | |
|---|---|---|
| Marin Wastewater Treatment Plant | | |
| Silicon Valley Clean Water | CA | 39.1666667 |
| Soscol Water Recycling Facility | CA | 1.7699115 |
| South County Regional Wastewater Authority | CA | 3.39622642 |
| Southeast San Francisco | CA | 56.3218391 |
| Turlock Regional Water Quality Control Facility | CA | 4.38596491 |
| UC Davis | CA | 0 |
| Vallejo Flood and Wastewater District Wastewater Treatment Plant | CA | 13.0434783 |
| Valley Sanitary District | CA | 16.6666667 |
| West County Wastewater District | CA | 1.03092784 |
| Windsor Wastewater Treatment, Reclamation, and Disposal Facility | CA | 0 |
| Winters - East Street Pump Station | CA | 0 |
| Woodland Water Pollution Control Facility | CA | 1.06382979 |
| Parker Water and Sanitation District North Water Reclamation Facility | CO | 1.88679245 |
| Parker Water and Sanitation District South Water Reclamation Facility | CO | 3.77358491 |
| City of Stamford, Water Pollution Control Authority | CT | 8.33333333 |
| Blue Plains Advanced Wastewater Treatment Plant | DC | 27.1604938 |
| Seaford Wastewater Treatment Facility | DE | 2.63157895 |

| | | |
|---|---|---|
| Altamonte Springs Regional Water Reclamation Facility | FL | 15.5963303 |
| Eastern Water Reclamation Facility | FL | 19.3548387 |
| Hamlin Water Reclamation Facility | FL | 7.77777778 |
| Loxahatchee River Environmental Control District | FL | 22.1238938 |
| MDWASD Central District WWTP | FL | 57 |
| MDWASD North District WWTF | FL | 65.7142857 |
| MDWASD South District WWTF | FL | 16.2790698 |
| Northeast Water Reclamation Facility | FL | 0 |
| Northwest Water Reclamation Facility | FL | 1.99004975 |
| South Water Reclamation Facility | FL | 23.4042553 |
| Southwest Water Reclamation Facility | FL | 6.14035088 |
| TPSmith Water Reclamation Facility | FL | 0.90909091 |
| Big Creek Water Reclamation Facility | GA | 9.64912281 |
| Camp Creek Water Reclamation Facility | GA | 3.84615385 |
| Johns Creek Environmental Campus | GA | 0 |
| Little River Water Reclamation Facility | GA | 11.4035088 |
| RM Clayton Water Reclamation Center | GA | 26.3736264 |
| South Columbus Water Resources Facility | GA | 6.89655172 |
| South River Water Reclamation Center | GA | 5.49450549 |

| | | |
|---|---|---|
| Utoy Creek Water Reclamation Center | GA | 0 |
| Hilo Wastewater Treatment Plant | HI | 0 |
| Honouliuli Wastewater Treatment Plant | HI | 0 |
| Kailua Regional Wastewater Treatment Plant | HI | 1.75438596 |
| Sand Island Wastewater Treatment Plant | HI | 3.50877193 |
| Wahiawa Wastewater Treatment Plant | HI | 0.89285714 |
| Waianae Wastewater Treatment Plant | HI | 0 |
| City of Clinton | IA | 0 |
| City of Marshalltown Water Pollution Control Plant | IA | 0.91743119 |
| Coralville Wastewater Treatment Facility | IA | 1.7699115 |
| Muscatine STP | IA | 0 |
| Ottumwa WPCF | IA | 0.87719298 |
| City of Coeur d'Alene Water Resource Recovery Facility | ID | 1.03092784 |
| Lander Street Water Renewal Facility | ID | 0.88495575 |
| West Boise Water Renewal Facility | ID | 3.57142857 |
| Glenbard Wastewater Authority | IL | 1.85185185 |
| Wheaton Sanitary District | IL | 4.62962963 |
| City of Carmel WWTP | IN | 5.26315789 |
| City of South Bend Wastewater Treatment Plant | IN | 10.7142857 |
| Dillman Road WWTP | IN | 4.03225806 |

| | | |
|---|---|---|
| Jeffersonville Downtown WWTP | IN | 0 |
| North Water Reclamation Facility | IN | 0.88495575 |
| Kansas City Treatment Plant #20 | KS | 3.33333333 |
| Lawrence Kansas River Wastewater Treatment Facility | KS | 85.5855856 |
| Municipal Wastewater Treatment Plant No. 1 (Kaw Point) | KS | 25 |
| Salina Wastewater Treatment Plant | KS | 49.122807 |
| Wolcott Wastewater Treatment Facility | KS | 31.0679612 |
| Morris Forman Water Quality Treatment Center | KY | 27.0833333 |
| SWBNO East Bank Wastewater Treatment Plant | LA | 13.5802469 |
| SWBNO West Bank Wastewater Treatment Plant | LA | 23.75 |
| Deer Island Treatment Plant | MA | 88.4955752 |
| Upper Blackstone Clean Water | MA | 2.91262136 |
| Hagerstown Wastewater Treatment Plant | MD | 0.87719298 |
| Marlay Taylor Water Reclamation Facility | MD | 0 |
| Brunswick Sewer District | ME | 9.70873786 |
| City of Bangor Wastewater Treatment Plant | ME | 8.41121495 |
| Lewiston Auburn Water Pollution Control Authority | ME | 67.3469388 |
| Portland Water District (East End Wastewater Treatment Facility) | ME | 95.1807229 |
| York Sewer District | ME | 10 |
| City of Ann Arbor Wastewater Treatment Plant | MI | 18.5840708 |

| | | |
|---|---|---|
| City of Warren Wastewater Treatment Plant | MI | 1.05263158 |
| Grandville Clean Water Plant | MI | 0 |
| Jackson Wastewater Treatment Plant | MI | 0 |
| Mt. Pleasant WRRF | MI | 1.86915888 |
| Traverse City Regional Waste Water Treatment Plant | MI | 25.4545455 |
| City of Mankato Water Resource Recovery Facility (WRRF) | MN | 20.3539823 |
| City Of Rochester MN Water Reclamation Plant | MN | 2.72727273 |
| Red Wing Wastewater Treatment Facility | MN | 0.88495575 |
| St. Cloud Nutrient, Energy and Water Recovery Facility | MN | 1.75438596 |
| 2C-Gautier POTW | MS | 0 |
| 7 C- Pascagoula Moss Point POTW | MS | 0 |
| Archie Elledge WWTP | NC | 16.3461538 |
| City of Wilson - Hominy Creek Water Reclamation Facility | NC | 0.95238095 |
| Johnnie Mosley Regional Water Reclamation Facility | NC | 0.95238095 |
| Northeast Water Resource Recovery Facility | NE | 0 |
| Theresa Street Water Resource Recovery Facility | NE | 0.87719298 |
| City of Dover Wastewater Treatment Facility | NH | 0 |
| Hall Street Wastewater Treatment Plant | NH | 12.2641509 |
| Penacook Wastewater Treatment Facility | NH | 0 |

| | | |
|---|---|---|
| Bayshore Regional Sewerage Authority | NJ | 0.87719298 |
| Cumberland County Utilities Authority | NJ | 8.73786408 |
| Passaic Valley Sewerage Commission | NJ | 57.9545455 |
| South Monmouth Regional Sewerage Authority | NJ | 10.5263158 |
| The Somerset Raritan Valley Sewerage Authority | NJ | 0 |
| Township of Ocean Sewerage Authority | NJ | 0 |
| Clark County Water Reclamation District (CCWRD) Flamingo Water Resource Center (FWRC) | NV | 18.2608696 |
| City of Oswego Wastewater Treatment Plant | NY | 0.89285714 |
| Ithaca Area Wastewater Treatment Facility | NY | 7.86516854 |
| Akron Water Reclamation Facility | OH | 6.48148148 |
| City of Youngstown Wastewater Treatment Plant | OH | 0 |
| Capital Region Water AWTF | PA | 40.3508772 |
| DELCORA Western Regional Treatment Plant | PA | 53.7634409 |
| University Park Water Reclamation Plant | PA | 0 |
| City of Yankton Wastewater Treatment Facility | SD | 0.88495575 |
| M.C. Stiles Wastewater Treatment Facility | TN | 32.0754717 |
| Moccasin Bend WWTP | TN | 23.364486 |
| City of Gainesville Wastewater Treatment Plant | TX | 0 |

| | | |
|---|---|---|
| City of Garland Rowlett Creek WWTP | TX | 4.76190476 |
| DCWT Dallas | TX | 23.5849057 |
| DCWT White Rock | TX | 28.9719626 |
| Duck Creek Wastewater Treatment Plant | TX | 24.137931 |
| Hollywood Road WWTP | TX | 1.7699115 |
| River Road WWTP | TX | 3.41880342 |
| SJRA WWTF No.1 | TX | 2.63157895 |
| SJRA WWTF No.2 | TX | 1.7699115 |
| SJRA WWTF No.3 | TX | 0 |
| South Laredo WWTP | TX | 13.7254902 |
| Southside Wastewater Treatment Plant (City of Dallas) | TX | 27.7777778 |
| Wichita Falls Resource Recovery Facility | TX | 2.72727273 |
| Zacate Creek WWTP | TX | 37.254902 |
| Central Valley Water Reclamation Facility | UT | 52.6785714 |
| Provo City Water Reclamation Facility | UT | 5.40540541 |
| Aquia Wastewater Treatment Facility | VA | 4.46428571 |
| Little Falls Run Wastewater Treatment Facility | VA | 16.0714286 |
| Town of Hillsville Wastewater Treatment Plant | VA | 20.7207207 |
| City of Essex Junction Wastewater Treatment Facility | VT | 0 |
| Montpelier Water Resource Recovery Facility | VT | 0.91743119 |
| South Burlington-Airport Parkway | VT | 0 |

| | | |
|---|---|---|
| WWTF | | |
| City of Snohomish Wastewater Treatment Plant | WA | 0 |
| Wausau Waterworks Wastewater Treatment Facility | WI | 5.30973451 |
| City of Wheeling, Water Pollution Control Division | WV | 3.77358491 |
